# Influences of Nutri-Grade front-of-pack labels on the consumption of sugar-sweetened and artificially sweetened beverages: moderating roles of the food environment and age

**DOI:** 10.64898/2025.12.14.25342245

**Authors:** Chia-Wen Wang, Mary Foong-Fong Chong, Pei Ma, Borame Lee Dickens, Yiyun Shou

## Abstract

Front-of-pack nutrition labels (FOPLs) have been adopted as a key government strategy to address the significant burden of diet-related noncommunicable diseases. However, research on public knowledge and perceptions of FOPLs and their relationships with sugar-sweetened beverage (SSB) and artificially sweetened beverage (ASB) consumption remains limited. A cross-sectional study with 2870 individuals was conducted to explore their knowledge and perceptions of Nutri-Grade, a national front-of-pack nutrition labeling scheme introduced in Singapore in 2022. Knowledge was not significantly associated with SSB consumption; however, individuals who perceived these labels more positively were significantly less likely to consume SSBs daily (AOR=0.72, *p*<0.001) and consumed fewer types of SSBs weekly (IRR=0.91, *p*<0.001). Moderation analyses indicated that greater knowledge of Nutri-Grade FOPLs was associated with a higher likelihood of SSB consumption among younger individuals and those residing in areas with higher house prices. Additionally, individuals with positive perceptions were more likely to consume SSBs daily when living in areas with food court density exceeding 2.99 per km² (AOR = 1.12, *p* = 0.004). Individuals with positive perceptions of Nutri-Grade FOPL were also less likely to consume ASBs (AOR=0.69, *p<*0.001), whereas, in contrast to SSBs, greater knowledge of Nutri-Grade FOPLs was associated with increased ASB consumption (AOR=1.42, *p*<0.001). These findings indicate that informational labeling itself is insufficient to change consumption behavior. Although positive perceptions of Nutri-Grade FOPLs provide a protective effect, this effect diminishes when the food environment is taken into account.

## 1 Introduction

The consumption of sugar-sweetened beverages (SSBs) is a key risk factor for obesity, type 2 diabetes (T2D), cardiovascular diseases (CVD), and certain cancers (Lara-Castor et al., 2025; Malik & Hu, 2022; Neelakantan et al., 2022; Qin et al., 2020a). In 2020, SSB consumption was estimated to be responsible for 2.2 million new T2D cases and 1.2 million new CVD cases worldwide (Lara-Castor et al., 2025). In response to the increasing burden of non-communicable diseases (NCDs), the World Health Organization has proposed that front-of-pack nutrition labels (FOPLs) be used as a cost-effective intervention (“Best Buy”) for preventing diet-related NCDs globally (World Health Organization, 2017). Evidence from a comprehensive meta-analysis of 221 studies further demonstrates that FOPLs effectively improve dietary outcomes at the population level (Kelly et al., 2024). FOPLs have been adopted as a prominent government strategy to promote healthier dietary behaviors worldwide. Notable examples are the Traffic-Light labeling system in the UK (Sacks et al., 2009), the Nutri-Score in EU countries (Julia et al., 2025), the Warning-label system in Chile (Taillie et al., 2020) and Peru (Lowery et al., 2022), and the Health Star Rating system in Australia (Brownbill et al., 2019), applied across a range of food and beverage products.

In response to the significant burden and risk of diabetes, Singapore, a developed country in Southeast Asia, was the first country in the world to introduce a mandatory FOPL specifically for SSBs; this policy became effective on December 30, 2022 (Ministry of Health of Singapore, 2021). Unlike most FOPL systems in other countries, which apply to both foods and beverages, Singapore’s Nutri-Grade FOPL system (see Figure 1) is distinctive in that it exclusively targets beverages, particularly SSBs, rather than food products broadly (Health Promotion Board, 2024). This focus recognizes that SSBs are a major, modifiable source of added sugar consumption [i.e., sugars added during food or beverage processing, such as sucrose and corn syrup (U.S. Food & Drug Administration, 2024)] and a key driver of overall sugar consumption in Singapore. Singaporeans consume 56 grams of sugar daily, surpassing the WHO’s recommended limit of 50 grams, with SSBs accounting for 52% of this total (Health Promotion Board, 2022b). A meta-analysis in Asian populations found that higher consumption of SSBs was associated with a 51% increased risk of T2DM (Neelakantan et al., 2022), higher than the corresponding risks reported in the US (15%) (Qin et al., 2020b) and in European countries (22%) (The InterAct consortium, 2013). This vulnerability is exacerbated by a rapid transition from traditional high-carbohydrate, low-fat diets to Western diets high in saturated fats, sugar, and salt over the past decade (Azzam, 2021; Bishwajit, 2015), including the rising popularity of freshly prepared drinks, such as bubble tea.

**Figure 1.**
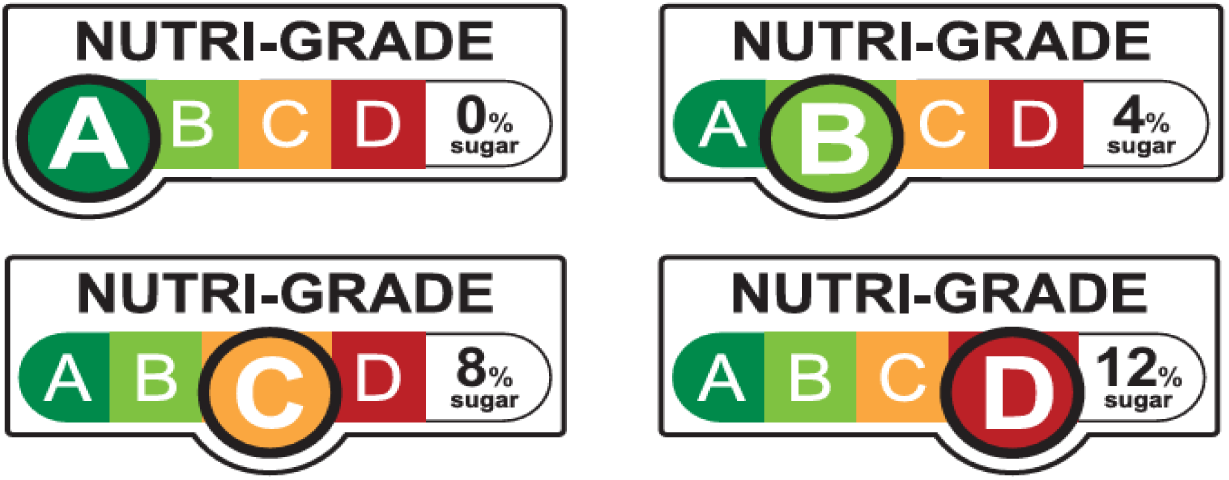
Nutri-Grade Mark on SSBs in Singapore.

FOPLs are designed to guide individuals in making informed decisions and selecting healthier options. However, their effectiveness depends largely on how people understand, perceive, and interpret the labels. Without a clear understanding of public knowledge and perceptions regarding FOPLs, it is difficult to evaluate whether such policies achieve their intended goal of reducing SSBs. This study aims to examine the links between the public’s knowledge and perceptions of Nutri-Grade FOPLs and the consumption of SSBs, 2 years after the roll-out of this novel policy initiative. We hypothesize that better knowledge and more positive perceptions of Nutri-Grade FOPLs would be associated with lower SSB consumption.

In addition, this study will investigate how built environment and age may moderate knowledge and perceptions of FOPLs and SSB consumption. Built environment that reflect neighborhood characteristics are known to shape SSB consumption behaviors (Duran et al., 2016). Understanding population heterogeneity in responding to FOPLs is essential for capturing the gaps in the FOPLs policy implementation.

Furthermore, it is observed that industries have begun to reformulate their beverage products by including the use of artificial sweeteners, so as to meet more favorable grades under the Nutri-Grade (e.g., B). Whether this has influenced ASB consumption is unclear. Therefore, this study also seeks to understand whether knowledge of and perception toward FOPLs are associated with ASB consumption.

Overall, this study primarily aims to examine the influences of knowledge and perception of Nutri-Grade FOPLs on SSB and ASB consumptions, even after taking into account other socio-ecological factors (environmental influences and individual knowledge and beliefs) that may influence beverage consumption (Fox & Timmer, 2020; McCormick et al., 2021; Wang et al., 2022). A secondary aim is to examine whether the food environment and age moderate the relationship between knowledge and perception of Nutri-Grade FOPLs on SSB consumption.

## 2 Methods

### 2.1 Study design and population

A cross-sectional study targeting adults aged 21 and above, with 2870 participants, was conducted in Singapore from February to April 2025. Participants were eligible to take part in this survey if they met the following criteria: (1) were Singaporean citizens or permanent residents; (2) were aged 21 years or older; (3) had no cognitive impairment; (4) provided consent to participate in the survey; (5) were able to read and understand English, Chinese, or Malay; and (6) resided in one of the sampling regions.

The survey was administered through the Qualtrics online survey platform. Data were collected through Pureprofile’s survey panel via a hybrid approach. Responses from 2310 participants were collected via online survey, and an additional 560 in-person surveys (e.g., street intercepts) were conducted to achieve the sampling quota, particularly those participants aged 50 or older. To ensure the representativeness of the sample, quota sampling was used to recruit participants while accounting for their gender, age, and ethnicity across five regions (north, northeast, east, central, and west) in Singapore. The two planning areas with the greatest number of residents were selected from each region. The sampling calculations were based on data from the Department of Statistics, Singapore, using the geographical distribution dashboard (Department of Statistics Singapore, 2025).

### 2.2 Measure

The detailed questionnaire is in Appendix A of the Supplementary Material.

#### 2.2.1 Demographic and health status

The participants self-reported their age, sex, ethnicity, education level, marital status, employment status, height, weight, and health conditions (e.g., mental health, diagnosed with chronic diseases, etc.).

#### 2.2.2 Influence of Nutri-Grade FOPL

##### Knowledge of Nutri-Grade labels

Information about the Nutri-Grade label is posted on the Health Promotion Board’s website to inform the public (Health Promotion Board, 2024). Participants’ knowledge of Nutri-Grade labels was assessed using eight items (e.g., “Nutri-Grade grades your drinks on the basis of their sugar and saturated fat levels”), with response options of correct, wrong, or don’t know. A total knowledge score was calculated for each participant based on the number of correctly answered questions, thus reflecting their level of knowledge about Nutri-Grade labels. In the current sample, the reliability of these eight items was acceptable ( α = .75 based on tetrachoric correlations).

##### Perception of Nutri-Grade labels

Participants’ perceptions of Nutri-Grade labels were assessed based on six items that captured clarity (e.g., “The Nutri-Grade label on sugar-sweetened beverages is easy for me to understand”), usefulness, informativeness, trustworthiness, reliability, and potential confusion when interpreting the Nutri-Grade labels. Responses were provided on a 7-point scale ranging from 1=strongly disagree to 7=strongly agree. Items reflecting negative perceptions were reverse-coded, so that higher scores indicated a more positive perception of Nutri-Grade labels. See Appendix A for the detailed items. In the current sample, the reliability of these six items was deemed acceptable (α = 79).

#### 2.2.3 Socio-ecological factors: Environmental influences

##### Built environment

The variables explored represent the local heterogeneous influence of the built environment, including socioeconomic and beverage exposure factors where values were estimated for each of the 16 postal code zones the participants resided in. For socioeconomic status, we calculated the mean house price for each zone, utilizing 2021-2024 Housing Development Board (HDB)^1^ resale flat prices as house price and housing type are associated with household income, education level, and overall socioeconomic status, serving as a proxy for neighborhood affluence.

We also calculated the proportion of commercial land use and the densities of food courts, cafés, bars, vending machines, and juice bars within the same 16 zones. The proportion of commercial land was derived from the Singapore Master Plan 2019 Land Use Layer, reflecting the degree of commercial development and overall bulk availability of beverage outlets. Further food and beverage outlet data were compiled from OpenStreetMap via QuickOSM queries and validated using the Singapore OneMap API. We calculated the outlet densities (number of outlets per km^2^ unit land area) for food courts (known locally as hawker centers), bars, cafes, vending machines, and juice bars.

##### Physical environment

Participants were asked to indicate the frequency of SSB availability in their household by responding to the following question: “How often are SSBs available in your home/household?” and “How often do you purchase SSBs via online delivery? (e.g., Foodpanda, Grab, FairPrice, Lazada, etc.)?” The response options for both questions included every day, every other day, every week, every other week, every month, and very rarely.

##### Social environment

Perceived social norms regarding acceptance of SSB consumption were measured using three items adapted from Phipps et al. (2020) (e.g., “Most people who are important to me would want me to drink sugar-sweetened beverages in a typical week”). The responses were rated on a 7-point scale ranging from 1=strongly disagree to 7=strongly agree. In the current sample, the three items exhibited acceptable reliability (α = .79). The exposure to SSB advertising was adapted from a previous study (Cervi et al., 2017). Participants’ susceptibility to SSB advertising was assessed using three items (e.g., “I want to try the advertised sugar-sweetened beverages”), in which context responses were scored on a 7-point scale ranging from 1=strongly disagree to 7=strongly agree.

#### 2.2.4 Socio-ecological factors: Individual knowledge and beliefs

##### Knowledge of SSBs and their impact on health

Participants’ knowledge of SSB on health impacts was measured using ten items (e.g., “Excess sugar-sweetened beverage consumption can contribute to increasing the risk of diabetes”), with response options of correct, wrong, and do not know. This scale was adapted from a previous study and from information provided on the Health Promotion Board’s website (Health Promotion Board, 2022a; Wang & Chen, 2022). A total knowledge score was calculated for each participant on the basis of the number of correct responses, thus reflecting their overall understanding of SSBs and the impact of SSBs on their health. A higher score indicates greater knowledge of SSBs. The 10 items exhibited good reliability (α = 81 based on tetrachoric correlations) in this study.

##### Perceived control of SSB consumption

Perceived control was measured using three items adapted from Collado-Rivera et al. (2018)(e.g., “If I wanted to, it would be easy for me to not drink sugar-sweetened beverages in a typical week.” Responses were scored on a 7-point scale ranging from 1=strongly disagree to 7=strongly agree. A higher score suggests that participants perceive greater control over their SSB consumption. The three items exhibited good reliability (α = 89).

##### Self-efficacy

The self-efficacy scale was adapted from a previous study by Wang and Chen (2022). Participants rated their confidence in reducing SSB consumption across six different contexts (e.g., “I am confident I am able to not drink sugar-sweetened beverages when I am tired.” The responses were scored on a 7-point scale ranging from 1=certain I could not to 7=certain I could. The scale exhibited excellent reliability (α = 91).

#### 2.2.5 SSB consumption

The categories of SSB consumption were adapted from the WHO’s 2023 Global Report on the Use of SSB Taxes (World Health Organization, 2023) and were tailored to reflect the local context of Singapore. Participants were asked to self-report their SSB consumption over a typical week. Before this, they were presented with a brief explanation of what SSBs are. They were then asked to report on their consumption of 11 types of SSBs: (1) sugar-sweetened carbonated beverages with sugar (e.g., Coca-Cola), (2) sugar-sweetened noncarbonated drinks (e.g., lemon water), (3) sugar-sweetened energy drinks (e.g., Red Bull), (4) sugar-sweetened sports/isotonic drinks (e.g., Pocari Sweat), (5) sweetened fruit drinks (e.g., apple juice), (6) sugar-sweetened malted drinks (e.g., Milo), (7) sugar-sweetened ready-to-drink (freshly prepared) tea (e.g., bubble tea), (8) sugar-sweetened ready-to-drink (freshly prepared) coffee (e.g., Kopi), (9) sugar-sweetened canned coffee or tea drinks (with or without milk) (e.g., Nescafe), (10) sugar-sweetened milk drinks (e.g., soya milk with sugar), and (11) traditional Chinese herbal sweetened beverages (e.g., Red Date Tea).

They were asked to report the frequency of their SSB consumption according to the following question: “In a typical week, how often do you consume the following drinks?” Response options included “none,” “1-3 times/week,” “4-6 times/week,” “1 time/per day,” “2 times/per day,” “3 or more times per day.” To estimate daily consumption, reported frequencies were converted to a continuous scale by dividing the weekly frequency by seven. For responses indicating 3 or more times per day, a maximum value of 3 was assigned. For instance, the midpoint of the “1–3 times/week” category was set at 2, which was then divided by 7 to yield an estimated daily consumption of 0.29 times per day. Daily consumption values across all SSB types were summed to calculate each participant’s total daily SSB consumption. Based on the distribution of participants’ responses, daily SSB consumption was further recategorized into five consumption groups: “no consumption,” “>0≤1 time/day,” “>1≤3 times/day,” “>3≤6 times/day,” and “>6 times/day.”

Additionally, the total number of different types of SSBs consumed by each participant per week was calculated. Each of the 11 SSB types was assigned a binary score: a response of “none” was coded as 0, while any reported frequency (“1–3 times/week,” “4–6 times/week,” “1 time/per day,” “2 times/per day,” or “3 or more times per day”) was coded as 1. The total score was obtained by summing all 11 beverage types, yielding a value ranging from 0 to 11, which reflected the diversity of SSB types consumed.

#### 2.2.6 ASB consumption

Participants were first provided with a brief explanation of ASBs to ensure an understanding of the term. They were subsequently asked to report their frequency of ASB consumption during a typical week by responding to the following question: “In a typical week, how often do you consume the following artificially sweetened beverages?” The items listed included (1) artificially sweetened carbonated beverages (e.g., Coke Zero), (2) artificial-sweetened noncarbonated drinks (e.g., zero-sugar-ice lemon tea), and (3) artificial sweetened energy drinks (e.g., Red Bull Zero). The same response options used for SSBs were applied, i.e., “none”, “1–3 times/week”, “4–6 times/week”, “1 time/per day”, “2 times/per day”, and “3 or more times per day.” Weekly frequencies were converted into estimated daily consumption using the same method as described for SSBs. The daily values for each ASB category were then summed up to generate the total daily ASB consumption.

### 2.3 Statistical analysis

Data analysis for this study was conducted using R (version 4.5.1) (Team, 2021) and R Studio. Demographic and health status variables were recoded for analysis: age was grouped as “21-39” (reference group), “40-59,” and “60 or above.” Sex was coded as “male” (reference group) and “female.” Ethnicity was coded as “Chinese” (reference group), “Malay,” “Indian,” and “others.” Education was collapsed into “below university” (reference group) and “university or above.” Marital status was categorized as “married with children” (reference group), “married without children,” “single,” and “widowed/divorced/separated.” Employment status was coded as “employed full-time” (reference group), “employed part-time,” “retired/homemaker,” “self-employed,” “unemployed,” and “others.” Self-reported height and weight values were calculated in terms of participants’ body mass index (BMI). BMI was categorized as “normal (18.5-22.9)” (reference group), “underweight (<18.5),” “overweight (23-27.4),” and “obese (≥27.5),” based on Asian-specific BMI cut-off points for adults in Singapore. Medical conditions were coded as “Yes” (reference group) and “No”.

This study examined three dependent variables: (1) total daily consumption of SSBs, (2) total number of different types of SSB consumed per week, and (3) total daily consumption of ASBs. The demographic variables and health status mentioned above were treated as control variables in statistical analyses.

Mixed-effects ordinal regression models were used to identify factors associated with the total daily consumption of SSBs and ASBs. Generalized linear mixed models using Template Model Builder were employed to examine factors associated with the total number of different types of SSB consumed per week. Models account for random effects by including postal code as a random intercept, thus facilitating a more robust estimation while controlling for the clustering of individuals residing in different geographical areas. The overall model fit was assessed using the Akaike information criterion (AIC).

Moderation analysis was conducted to examine whether the associations between knowledge and perception of Nutri-Grade labels and SSB consumption varied across built environment and age groups, while controlling demographic variables, health status, and socio-ecological factors.

## 3 Results

In total, 2,870 participants completed the survey and were included in the analysis. The sociodemographic characteristics of the participants are presented in Table 1. The mean age of the participants was 45.52 years (SD=12.71). Among them, 51.43% were female, 71.05% were Chinese, 46.06% held a university degree or above, 58.95% were married with children, and 72.09% were employed on a full-time basis. Furthermore, 37.77% were classified as overweight, 11.78% as obese, and 31% had been diagnosed with at least one medical condition (e.g., diabetes or hypertension).

**Table 1.**
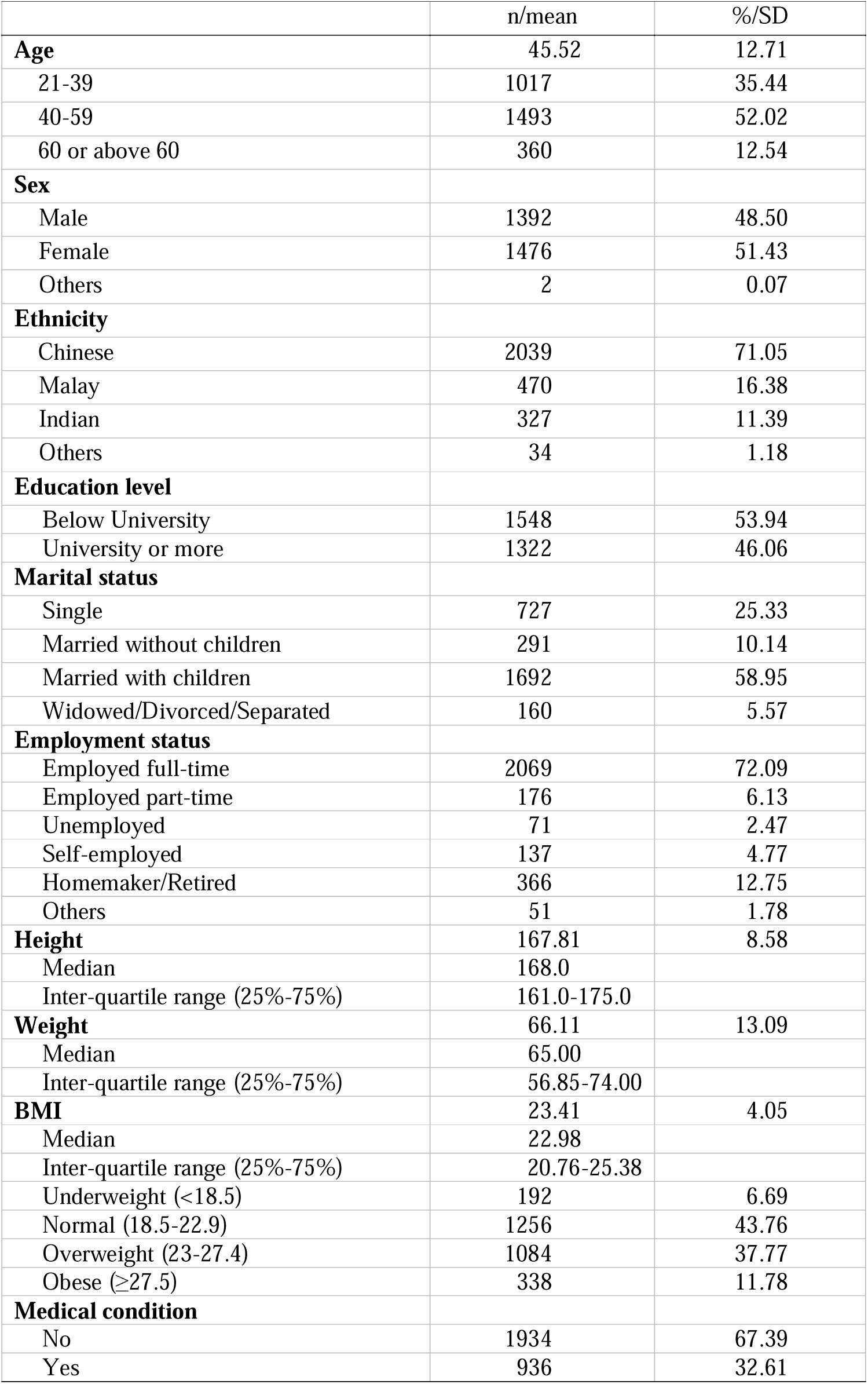
Socio-demographic characteristics of the participants (n=2,870)

The Spearman correlation matrix (Figure 2) illustrates the relationships among the three dependent variables and key predictors. Notably, strong correlations among total daily consumption of SSBs, total number of different types of SSB consumed, and ASB consumption were revealed, suggesting that higher overall consumption tends to co-occur across these beverage categories. Additionally, a higher frequency of SSB consumption and the total number of different types of SSB consumed were positively associated with household availability of SSBs, purchase of SSBs via online delivery, perceived social norm of acceptance of SSB consumption, exposure to SSB advertising, and knowledge of Nutri-Grade FOPLs. In contrast, total SSB consumption and total number of different types of SSB consumption were negatively correlated with age, perceived control over SSB consumption, self-efficacy, and perceptions of Nutri-Grade FOPLs. Regarding ASB consumption, it was positively associated with knowledge of SSBs and their impact on health, household availability of SSBs, purchase of SSBs via online delivery, perceived social norm of acceptance of SSB consumption, exposure to SSB advertising, and knowledge of Nutri-Grade FOPLs. Conversely, it was negatively associated with age, perceived control over SSB consumption, self-efficacy, and perceptions of Nutri-Grade FOPLs.

**Figure. 2.**
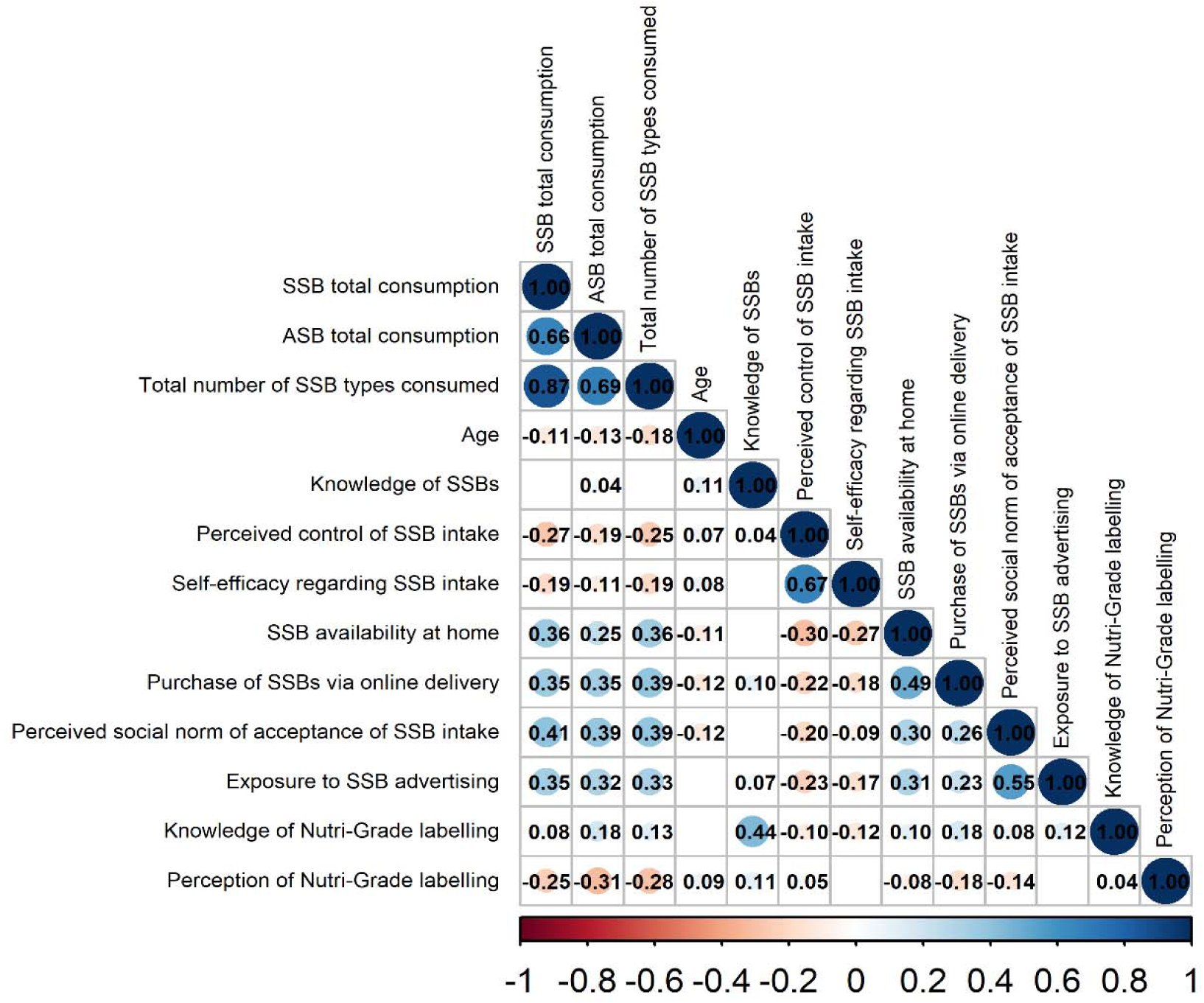
The Spearman correlation matrix of the dependent variables and other key variables. Blank cells represent correlations that are not statistically significant.

### 3.1 FOPLs Knowledge and Perceptions on SSB and ASB Consumption

Upon adjusting for demographic variables and health status, the findings suggest that higher knowledge levels with respect to Nutri-Grade FOPLs were no longer associated with likelihood of consuming SSBs daily (AOR=1.04, 95% CI: 0.96–1.13, *p*=0.354) or the number of types of SSBs weekly (IRR=1.02, 95% CI: 1.00–1.04, *p*=0.115). Additionally, individuals with positive perceptions of Nutri-Grade FOPLs were less likely to consume SSBs daily (AOR=0.72, 95% CI: 0.67–0.78, *p*<0.001) and fewer types of SSBs weekly (IRR=0.91, 95% CI: 0.89–0.92, *p*<0.001) (Table 2).

**Table 2.**
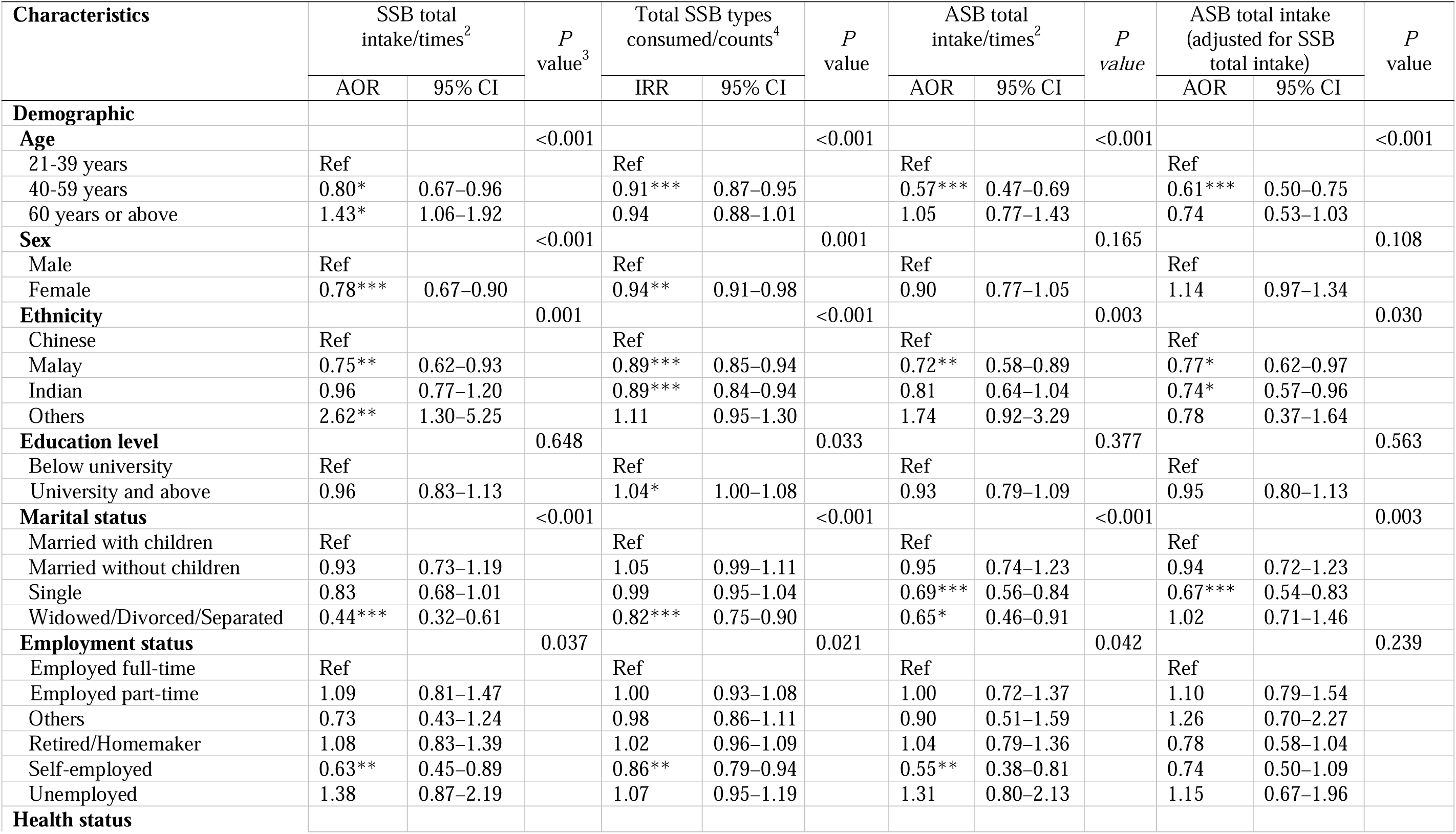
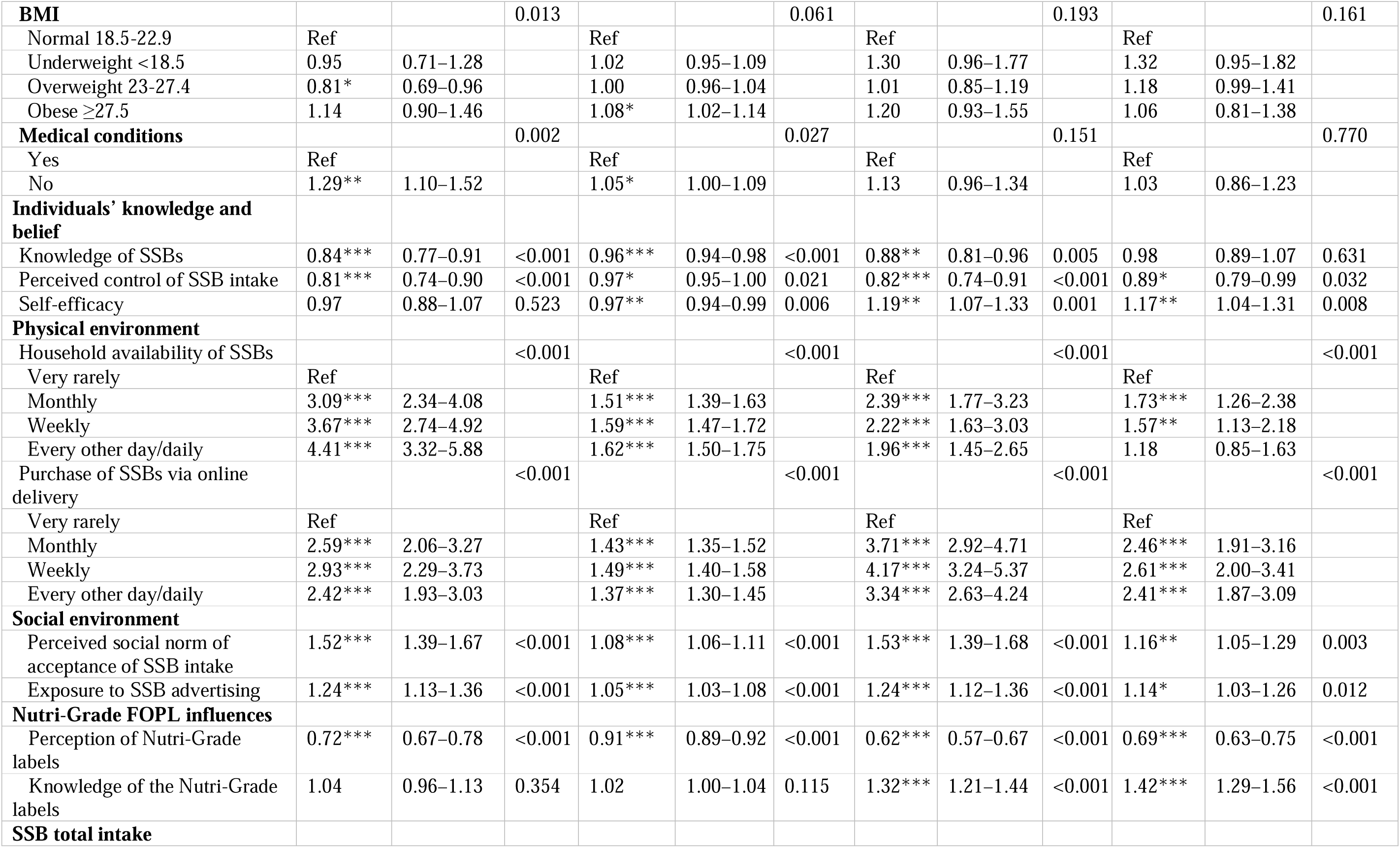
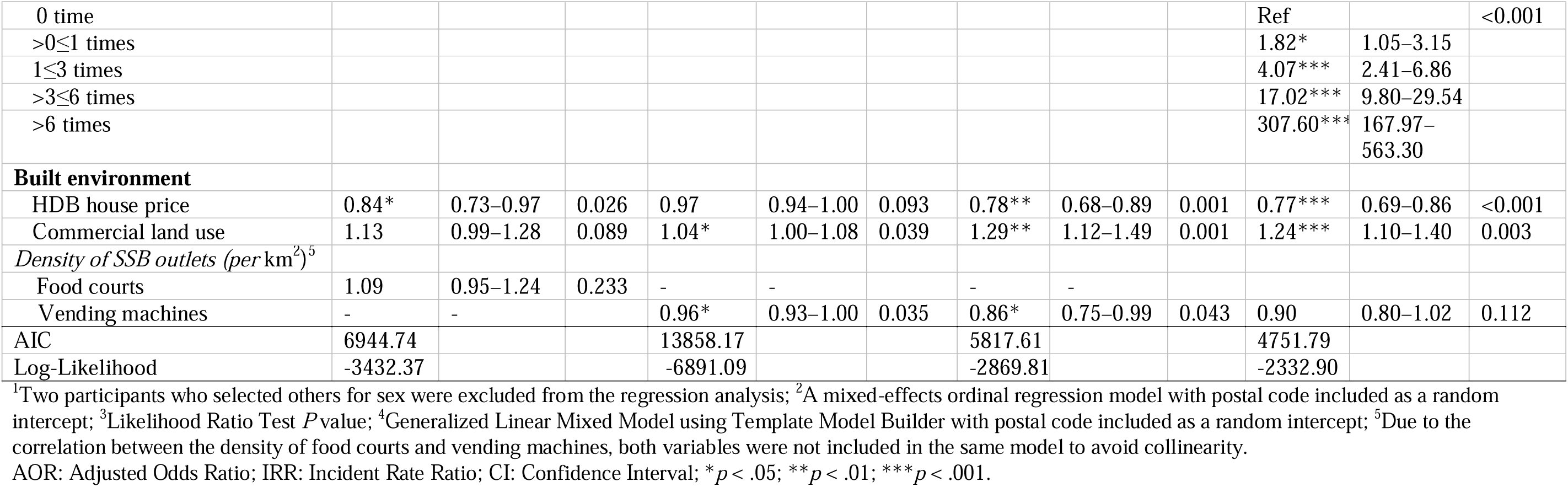
Factors associated with daily total consumption of SSBs, the total number of different types of SSBs consumed weekly, and daily consumption of ASBs in Singapore (n = 2,868)^1^.

Similarly, upon adjustment for demographic variables and health status, individuals with positive perceptions of Nutri-Grade FOPLs were less likely to consume ASBs (AOR=0.69, 95% CI=0.63–0.75, *p<*0.001), whereas greater knowledge of Nutri-Grade FOPLs was associated with increased ASB consumption, even after adjusting for SSB consumption (AOR=1.42, 95% CI=1.29–1.56, *p*<0.001)(Table 2).

### 3.2 Moderation of the Built Environment

Upon adjusting for demographic variables and health status, the findings suggest that HDB house prices, the proportion of commercial land use, and the densities of bars, cafés, and vending machines significantly moderate the relationship between knowledge of Nutri-Grade FOPLs and daily consumption of SSBs. Specifically, greater knowledge of Nutri-Grade FOPLs was associated with higher SSB daily consumption among individuals residing in areas with higher HDB house prices (AOR=1.30, 95% CI: 1.21–1.41, *p*<0.001).

Similarly, greater knowledge of Nutri-Grade FOPLs was associated with a higher likelihood of daily SSB consumption among individuals living in areas with a higher proportion of commercial land use and greater densities of bars, cafes, and vending machines. However, after controlling for the most significant interaction (HDB house price × knowledge of the Nutri-Grade FOPLs), only the interaction between bar density and knowledge of the Nutri-Grade FOPLs remained significant (AOR=1.12, 95% CI: 1.03–1.23, *p*=0.011).

Likewise, greater knowledge of Nutri-Grade FOPLs was associated with a higher likelihood of SSB types consumed weekly among individuals residing in areas with higher HDB house prices and greater bar density (HDB house price: IRR=1.05, 95% CI: 1.03–1.07, *p*<0.001; bar density: IRR=1.03, 95% CI: 1.01–1.06, *p*=0.006).

In contrast, greater knowledge of Nutri-Grade FOPLs was associated with a reduced likelihood of SSB types consumed weekly among individuals living in areas with a higher density of juice bars (IRR=0.98, 95% CI=0.96–1.00, *p*=0.019). After controlling for the most significant interaction (HDB house price ×knowledge of the Nutri-Grade FOPLs), only juice bar density was significant (IRR=0.97, 95% CI=0.96–0.99, *p*= 0.007).

Furthermore, after adjusting for demographic variables and health status, individuals with positive perceptions of Nutri-Grade FOPLs were less likely to consume SSBs daily when residing in areas with food court density below 2.99 per km². In contrast, those with positive perceptions were more likely to consume SSBs daily when living in areas with food court density exceeding 2.99 per km² (AOR = 1.12, 95% CI = 1.04–1.21, p = 0.004).

This suggests that the protective effect of Nutri-Grade FOPL perception on SSB consumption was attenuated as food court density increased. However, no other built environment factors significantly moderate the relationship between the perception of Nutri-Grade FOPLs and SSB daily consumption and total number of SSB types consumed weekly (Table 3).

**Table 3.** Moderation effects of knowledge and perception of Nutri-Grade FOPL influences and built environment factors on SSB consumption in Singapore (n = 2,868)

| Interaction term<sup>1</sup> | SSB total intake daily |  |  |  | Total SSB types consumed weekly |  |  |  |
| --- | --- | --- | --- | --- | --- | --- | --- | --- |
|  | Knowledge of the Nutri-Grade FOPLs |  | Perception of Nutri-Grade FOPLs |  | Knowledge of the Nutri-Grade FOPLs |  | Perception of Nutri-Grade FOPLs |  |
|  | AOR | 95% CI | AOR | 95% CI | IRR | 95% CI | IRR | 95% CI |
| ×HDB house price | 1.30*** | 1.21–1.41 | 1.00 | 0.93–1.08 | 1.05*** | 1.03–1.07 | 0.99 | 0.97–1.01 |
| ×Commercial land use | 1.09* | 1.02–1.18 | 0.98 | 0.90–1.05 | 1.01 | 0.99–1.03 | 1.00 | 0.98–1.02 |
| ×Food Courts | 1.00 | 0.93–1.07 | 1.12** | 1.04–1.21 | 1.00 | 0.98–1.01 | 1.01 | 0.99–1.02 |
| ×Bars | 1.17*** | 1.07–1.27 | 0.97 | 0.89–1.06 | 1.03** | 1.01–1.06 | 1.00 | 0.98–1.02 |
| ×Cafes | 1.13** | 1.05–1.22 | 1.03 | 0.96–1.11 | 1.02 | 1.00–1.04 | 1.00 | 0.98–1.02 |
| ×Vending machines | 1.09* | 1.01–1.18 | 1.02 | 0.94–1.10 | 1.01 | 0.99–1.03 | 1.00 | 0.98–1.02 |
| ×Juice bars | 0.96 | 0.89–1.04 | 1.03 | 0.96–1.11 | 0.98* | 0.96–1.00 | 1.00 | 0.98–1.02 |
| ×age 40-59 (reference group: age 21-39) | 0.81* | 0.69–0.95 | 1.08 | 0.93–1.26 | 0.96 | 0.93–1.00 | 0.99 | 0.95–1.02 |
| ×age 60 or above (reference group: age 21-39) | 0.92 | 0.72–1.18 | 1.00 | 0.78–1.28 | 0.97 | 0.91–1.03 | 0.99 | 0.93–1.05 |

<sup>1</sup>This table presents only the interaction term results, whereas the full model estimates, adjusted for covariates, are reported in Table S1-S5 in Appendix B of the Supplementary Material.
AOR: Adjusted Odds Ratio; CI: Confidence Interval; \* $p < .05$ ; \*\* $p < .01$ ; \*\*\* $p < .001$ .

### 3.3 Moderation of age groups

Upon adjusting for demographic variables and health status, the results suggest that increased knowledge of Nutri-Grade FOPLs was associated with a lower likelihood of daily consumption of SSBs among those aged 40-59 compared to those aged 21 to 39 (AOR=0.81, 95% CI: 0.69–0.95, *p*=0.011). However, no such relationship was observed for those aged 60 or above (AOR=0.92, 95% CI: 0.72–1.18, *p*=0.523). Furthermore, the association between the positive perceptions of Nutri-Grade FOPLs and SSB daily consumption did not differ significantly across age groups (Table 3). Additionally, age did not moderate the associations between knowledge or perception of Nutri-Grade FOPLs and the total number of SSB types consumed weekly.

### 3.4 Individual and Social Factors Associated with SSBs

Upon adjusting for demographic variables and health status, the findings indicate that greater knowledge of SSBs and their impact on health was associated with lower daily SSB consumption and a reduced number of different types of SSBs consumed weekly. However, individuals with greater perceived control over SSB consumption reported both lower daily consumption and fewer types of SSBs consumed weekly. Conversely, while self-efficacy was not significantly associated with lower daily consumption, it was significantly associated with a reduced number of different types of SSBs consumed weekly.

Finally, greater availability of SSBs at home or through online delivery was associated with a greater likelihood of daily SSB consumption and a greater variety of SSBs weekly. Additionally, both perceived social acceptance of SSB consumption and exposure to SSB advertising were positively linked to higher daily consumption and an increased number of different types of SSB consumed weekly. These findings suggest that both physical and social environments play significant roles in influencing SSB consumption behaviors.

## 4 Discussion

This study suggests that greater knowledge of Nutri-Grade FOPLs was not significantly associated with SSB consumption but higher ASB consumption in the Singaporean residents. Moderation analyses indicated that greater knowledge of Nutri-Grade FOPLs was associated with a higher likelihood of SSB consumption among younger individuals and those residing in areas with higher house prices. Individuals who perceived these labels more positively reported significantly lower consumption of SSB, but this association is attenuated in areas with a high density of food courts. The findings that knowledge of Nutri-Grade FOPLs was not significantly associated with SSB consumption contrast with limited prior research, which found that the comprehension of nutrition labels was inversely associated with SSB consumption (Persoskie et al., 2017). The relatively recent introduction of Nutri-Grade and the potential disconnect between knowledge and perception may be limiting its current behavioral impact, even among those who are well informed about the label (Nayga Jr, 1999). The same individuals may be dependent on their high institutional trust in the government on public health knowledge, reducing the level of scrutiny or personal verification required before accepting guidance or messaging, resulting in positive perceptions of policy legitimacy even in the absence of detailed knowledge (Liu et al., 2015).

Moderation analyses indicated that greater knowledge of Nutri-Grade FOPLs was associated with a higher likelihood of SSB consumption among younger individuals and those residing in areas with higher house prices, used here as a proxy for socio-economic status (SES). One possible explanation for this is that young adults and individuals in affluent neighborhoods are likely to have a greater education level. They may be more attentive to Nutri-Grade FOPLs and have greater nutritional awareness and more accurate self-reporting among higher SES groups (Chen et al., 2012; Hendrie et al., 2008; Lahti-Koski et al., 2012).

The density of food courts also moderated the relationship between perception of Nutri-Grade and daily SSB consumption. Individuals who perceived the labels more positively were generally less likely to consume SSBs daily; however, this association was attenuated in areas where food court density exceeded 2.99 per km^2^. In Singapore, food courts and hawker centers play a central role in everyday life and are integral to the built environment.

Approximately 77% of Singaporeans reside in public housing estates (HDBs) (Department of Statistics Singapore, 2024), where such food outlets are embedded in neighborhood infrastructure. This urban configuration substantially enhances accessibility to SSBs by providing high convenience and short travel distances, reinforcing habitual consumption opportunities despite positive perceptions of Nutri-Grade labeling.

We found that individuals who perceived the labels more positively were significantly less likely to consume both SSBs and ASBs. Unlike the case of SSBs, we observed that greater knowledge of Nutri-Grade FOPLs was associated with higher ASB consumption, likely reflecting the widespread marketing of ASBs as being healthier substitutes and a “Healthier Choice”. It is important to note that the FOPL systems targeting SSBs rely on sugar content as their primary evaluation criteria. However, these standards may fail to adequately inform the public about artificial sweetener content and potentially mislead public perception regarding ASB healthfulness. Although the U.S. Food and Drug Administration has approved six artificial sweeteners (e.g., Aspartame, Acesulfame potassium, and Sucralose) as safe for the general population with established acceptable daily intake levels (U.S. Food & Drug Administration, n.d.), current evidence indicates that artificial sweeteners might not be a risk-free alternative to sugar over the long term. Several studies have demonstrated potential links between ASB intake and adverse health outcomes such as obesity, T2D, and CVD (Diaz et al., 2023; Huang et al., 2017; Ruanpeng et al., 2017). It is concerning that the current FOPL system targeting SSBs may inadvertently position ASBs as healthier alternatives due to their sugar-free formulation, thereby posing new public health challenges.

Similar behavioral patterns have been reported in the increasing use of e-cigarettes over traditional tobacco (Pesko et al., 2020), raising concerns about unintended health consequences (Jankowski et al., 2019). Similarly, a recent review has suggested that ASB consumption is associated with elevated risks of obesity and diabetes mellitus (Diaz et al., 2023). Individuals may feel that ASBs are harmless, thus increasing their consumption (Rebolledo et al., 2022), which could reduce overall sugar consumption but potentially introduce other risks, as the long-term effects of increased ASB consumption have yet to be ascertained. For this reason, multiple FOPLs have been updated to distinguish non-nutritive ASBs (Merz et al., 2024). ASB consumption may warrant similar community engagement efforts as SSBs, emphasizing moderation due to evidence linking excessive ASB consumption with increased risks of all-cause and cardiovascular mortality (Chen et al., 2024).

## 5 Conclusions

To summarize, no statistically significant association was found between knowledge of Nutri-Grade FOPL and SSB consumption, suggesting that social norms, more than awareness, govern SSB consumption behavior. Environmental and sociodemographic factors, such as age, SES, and accessibility of food outlets, could alter the effects of labeling. Rising ASB consumption, reflecting a shift towards perceived healthier SSB substitutes, and associated with increased knowledge of Nutri-Grade FOPL, warrants further investigation of its long-term impacts.

## 6 Policy implications

These findings indicate that informational labeling itself is insufficient to change consumption behavior and that effective sugar reduction labeling policies should account for the food environment. The absence of association between knowledge of Nutri-Grade FOPL and SSB consumption suggests that awareness does not necessarily translate into behavioral change when consumption is shaped by social norms and accessibility. Although positive perceptions of Nutri Grade were associated with lower consumption, this relationship was weakened in areas with greater densities of food outlets, showing that the physical environment can constrain individual intentions. Policy approaches should therefore extend beyond information provision to include policies that alter the choice environment, such as limiting the availability of SSB in high-exposure settings, improving access to unsweetened alternatives, and applying fiscal or placement strategies to encourage healthier options. The positive association between greater knowledge and higher consumption among younger and higher-income adults suggests a need for targeted communication that addresses social and generational drivers, including the influence of social media. The increase in ASB consumption, at least partially driven by FOPLs, highlights the need to refine labeling to better distinguish non-nutritive sweeteners and to clarify potential long-term health implications. Overall, the results support a coordinated approach that combines labeling, education, and environmental change to enable and sustain healthier beverage choices.

## Supporting information

Appendix A of the Supplementary Material

Appendix B of the Supplementary Material

## Data Availability

All data produced in the present study are available upon reasonable request to the authors

## Acknowledgements

The authors appreciate all the participants’ contributions to this study.

## Ethics statement

This study was approved by the Institutional Review Board of the National University of Singapore (NUS-IRB-2024-1018) and adhered to the Declaration of Helsinki. Informed consent was obtained from all participants for both their participation and the use of their study results.

## Data Availability Statement

Data are available upon reasonable request to the authors

## CRediT authorship contribution statement

**Chia-Wen Wang** Conceptualization, Data curation, Formal analysis, Funding acquisition, Investigation, Methodology, Project administration, Writing – original draft. **Mary Foong-Fong Chong:** Conceptualization, Investigation, Methodology, Project administration, Writing – review and editing. **Pei Ma:** Methodology; Writing – review and editing. **Borame Lee Dickens**: Conceptualization, Investigation, Methodology, Project administration, Supervision, Writing – review and editing.

**Yiyun Shou:** Conceptualization, Investigation, Methodology, Project administration, Supervision, Writing – review and editing.

## Footnotes

1 In Singapore, there are various housing types, including Housing & Development Board (HDB) flats, private condominiums, and landed properties. The majority of Singapore’s population (77.4%) resides in public housing, primarily managed by the HDB (Department of Statistics Singapore 2024).

