## Appendix A of the Supplementary Material for "Influences of Nutri-Grade front-of-pack labels on the consumption of sugar-sweetened and artificially sweetened beverages: moderating roles of the food environment and age"

**Part 1: Demographic and health information**

1. What is your residence status?

- Singaporean citizen
- Singapore Permanent Resident
- Other

2. What is your age?

_____________

3. How do you describe yourself?

- Male
- Female
- Non-binary/ third gender
- Prefer to self-describe: __________

4. Which racial group do you belong to?

- Chinese
- Malay
- Indian
- Other

5. What are the first two digits of your residential postal code?

££

6. What is the highest level of education you have completed/attained?

- Pre-primary
- Primary
- Secondary
- Post-secondary (non-tertiary): General and vocational (GCE ‘A’ Levels / Nitec certificate / Higher Nitec)
- Polytechnic diploma
- Professional qualifications and other diploma
- Bachelor’s or equivalent
- Postgraduate diploma/certificate
- Master’s and doctorate or equivalent
- Other (please specify): ____

7. What is your marital status?

- Single
- Married without children
- Married with children
- Widowed
- Divorced/Separated
- Other (please specify): ____

8. What is your employment status?

- Employed full-time
- Employed part-time
- Unemployed, seeking employment
- Unemployed, not seeking employment
- Self-employed
- Student
- Retired
- Homemaker
- Other (please specify): _______

9. What is your height in cm? (Please fill in up to one decimal place) _________
10. What is your weight in kg? (Please fill in up to one decimal place) __________
11. Have you ever been diagnosed with or suffered from one or more of the following conditions? (Select all that apply)

- Pre-diabetes
- Diabetes Mellitus
- Stroke
- Major Depression
- Osteoarthritis
- Anxiety
- Chronic Kidney Disease
- Rheumatoid Arthritis
- Gout
- Eating disorder
- Fatty liver
- Overweight
- Obesity
- Others, please specify: __________________
- No diagnosed medical condition

**Part 2: Knowledge and perception of the Nutri-Grade FOPL label**

12. Based on what you know, please answer the following questions.

|  |  | True | False | Don’t know |
| --- | --- | --- | --- | --- |
| (1) | Nutri-Grade “grades” your drinks based on their sugar and saturated fat levels. | - *(1)* | - *(0)* | - *(0)* |
| (2) | The Nutri-Grade mark can be found on both packaged drinks and menus or menu boards for freshly prepared drinks like kopi, teh, malted drinks, bubble tea, shakes, smoothies and more. | - *(1)* | - *(0)* | - *(0)* |
| (3) | Healthier choice drinks are Nutri-Grade C or D.  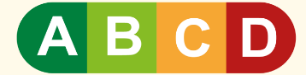 | - *(0)* | - *(1)* | - *(0)* |
| (4) | Some fruit juices can be graded “C” or “D”. | - *(1)* | - *(0)* | - *(0)* |
| (5) | Drinks graded “A” contain 0g of sugar and saturated fat per 100 ml. | - *(0)* | - *(1)* | - *(0)* |
| (6) | Some soft drinks are graded as “A” or “B”, while certain dairy drinks, like full-fat milk, are graded as “C” or “D”. This implies that soft drinks are healthier than these dairy drinks. | - *(0)* | - *(1)* | - *(0)* |
| (7) | The nutrition information for Coke Zero says that it contains 0g of total sugars and fat, it includes a permitted non-sugar substitute and therefore cannot be graded as "A". | - *(1)* | - *(0)* | - *(0)* |
| (8) | Advertisements of Nutri-Grade beverages graded “D”, are prohibited across all media platforms (e.g. broadcast, print, out-of-home, on-ground, online) except for advertisements of pre-packaged beverages at point-of-sale (POS) platforms within variety shops (e.g., supermarkets, convenience stores). | - *(1)* | - *(0)* | - *(0)* |

13. Regarding your perception of the Nutri-Grade label on sugar-sweetened beverages, to what extent do you agree or disagree with the following statements?

|  |  | 1 = Strongly Disagree | 2 = Disagree | 3 = Somewhat Disagree | 4 = Neutral | 5 = Somewhat Agree | 6 = Agree | 7 = Strongly Agree |
| --- | --- | --- | --- | --- | --- | --- | --- | --- |
| (1) | The Nutri-Grade label on sugar-sweetened beverages is **easy** for me to understand. |  |  |  |  |  |  |  |
| (2) | The Nutri-Grade label on sugar-sweetened beverages is **useless** for me. |  |  |  |  |  |  |  |
| (3) | The Nutri-Grade label on sugar-sweetened beverages is **informative** for me. |  |  |  |  |  |  |  |
| (4) | The Nutri-Grade label on sugar-sweetened beverages is **unreliable** for me. |  |  |  |  |  |  |  |
| (5) | The Nutri-Grade label on sugar-sweetened beverages is **trustful** for me. |  |  |  |  |  |  |  |
| (6) | The Nutri-Grade label is **confusing** when purchasing certain drinks (e.g. full cream milk). |  |  |  |  |  |  |  |

**Part 3: Environmental influences**

|  |  | Every day | Every other day | Every week | Every other week | Every month | Very rarely |
| --- | --- | --- | --- | --- | --- | --- | --- |
| (1) | available in your home/household? |  |  |  |  |  |  |
| (2) | are available for purchase via online delivery (e.g. foodpanda, Grab, FairPrice, Lazada, etc.)? |  |  |  |  |  |  |

14. How common are sugar-sweetened beverages…

|  |  | 1 = Strongly Disagree | 2 = Disagree | 3 = Somewhat Disagree | 4 = Neutral | 5 = Somewhat Agree | 6 = Agree | 7 = Strongly Agree |
| --- | --- | --- | --- | --- | --- | --- | --- | --- |
| (1) | I want to try the advertised sugar-sweetened beverages. |  |  |  |  |  |  |  |
| (2) | I think the advertised sugar-sweetened beverages will taste good. |  |  |  |  |  |  |  |
| (3) | I trust the advertised messages of sugar-sweetened beverages. |  |  |  |  |  |  |  |

15. When I see advertisements for sugar-sweetened beverages…
16. When you think about those people who are the most important to you (e.g. parents, siblings, spouse, colleagues, friends, teachers, etc.) ...To what extent do you agree or disagree with the following statements?

|  |  | 1 = Strongly Disagree | 2 = Disagree | 3 = Somewhat Disagree | 4 = Neutral | 5 = Somewhat Agree | 6 = Agree | 7 = Strongly Agree |
| --- | --- | --- | --- | --- | --- | --- | --- | --- |
| (1) | Most people who are important to me would **want me** to drink sugar-sweetened beverages in a typical week. |  |  |  |  |  |  |  |
| (2) | Most people who are important to me would **approve** of me drinking sugar-sweetened beverages in a typical week. |  |  |  |  |  |  |  |
| (3) | Most of the people who are important to me **drink** sugar-sweetened beverages in a typical week. |  |  |  |  |  |  |  |

**Part 4: Individual knowledge and belief**

17. Please indicate which statements are true or false to the best of your knowledge. *[Scores in brackets ()]*

|  |  | True | False | Don’t know |
| --- | --- | --- | --- | --- |
| (1) | Consuming too much sugar can contribute to obesity. | - *(1)* | - *(0)* | - *(0)* |
| (2) | Excess sugar-sweetened beverage consumption can contribute to increasing the risk of diabetes. | - *(1)* | - *(0)* | - *(0)* |
| (3) | Replacing white sugar with honey for the same level of sweetness substantially reduces the overall sugar intake. | - *(0)* | - *(1)* | - *(0)* |
| (4) | Excess sugar-sweetened beverage consumption could lead to bone loss. | - *(1)* | - *(0)* | - *(0)* |
| (5) | Drinking fresh fruit juices is as beneficial as eating fresh fruits in terms of nutritional value. | - *(0)* | - *(1)* | - *(0)* |
| (6) | Sugar that is found in 100% fruit juice is healthier than the sugar that is added to make soda. | - *(0)* | - *(1)* | - *(0)* |
| (7) | Brown sugar is a healthy alternative to white sugar. | - *(0)* | - *(1)* | - *(0)* |
| (8) | The sugar that is found naturally in fruits and milk has the same impact on tooth health as sugar that is added to foods and drinks when they are being prepared. | - *(0)* | - *(1)* | - *(0)* |
| (9) | Consuming sugar-sweetened beverages for hydration has the same effects on health as consuming water. | - *(0)* | - *(1)* | - *(0)* |
| (10) | The maximum recommended amount of added sugar for an average, healthy adult is 50 grams (about 10 teaspoons). | - *(1)* | - *(0)* | - *(0)* |

|  |  | 1 = Strongly Disagree | 2 = Disagree | 3 = Somewhat Disagree | 4 = Neutral | 5 = Somewhat Agree | 6 = Agree | 7 = Strongly Agree |
| --- | --- | --- | --- | --- | --- | --- | --- | --- |
| (1) | I am sure I am able to **not drink** sugar-sweetened beverages in a typical week. |  |  |  |  |  |  |  |
| (2) | If I wanted to, it would be easy for me to **not drink** sugar-sweetened beverages in a typical week. |  |  |  |  |  |  |  |
| (3) | I have complete control over **not drinking** sugar-sweetened beverages in a typical week. |  |  |  |  |  |  |  |

18. To what extent do you agree or disagree with the following statements?
19. I am *confident* I am able to not **drink** sugar-sweetened beverages …

|  |  | 1 = Certain I Could Not | 2 = Very Unlikely I Could | 3 = Somewhat Unlikely I Could | 4 = Neutral | 5 = Somewhat Likely I Could | 6 = Very Likely I Could | 7 = Certain I Could |
| --- | --- | --- | --- | --- | --- | --- | --- | --- |
| (1) | when I am tired. |  |  |  |  |  |  |  |
| (2) | when I am stressed. |  |  |  |  |  |  |  |
| (3) | when I am happy. |  |  |  |  |  |  |  |
| (4) | after work/exams/classes. |  |  |  |  |  |  |  |
| (5) | when I eat meals. |  |  |  |  |  |  |  |
| (6) | when gathering with family, friends, or classmates. |  |  |  |  |  |  |  |

Sugar-sweetened beverages are drinks that contain added sugars in various forms, such as brown sugar, corn syrup, glucose, fructose, high-fructose corn syrup, honey, maltose, molasses, raw sugar, and sucrose.

Examples of sugar-sweetened beverages include but are not limited to carbonated drinks with sugar (*e.g. Coca-Cola, Pepsi, 100 Plus, Fanta, 7up, etc.),* sweetened fruit drinks (*e.g. apple juice, mango juice, watermelon juice, etc.),* sugar-sweetened ready-to-drink coffee *(e.g. Kopi, Kopi O, Kopi C, Yuan Yang,* *Coffee Frappuccino, etc.), s*ugar-sweetened ready-to-drink tea *(e.g. Teh, Teh O, Teh Tarik, bubble tea, lemon tea, milk tea, etc.)*.

Artificially sweetened beverages, also known as non-nutritive sweetened beverages, are drinks that contain artificial sweeteners such as aspartame, acesulfame potassium (Ace-K), saccharin, sucralose, and others.

Examples of artificially sweetened beverages include, but are not limited to, *Coke Zero, 100 Plus Zero, Sprite Zero, Diet Coca-Cola, Diet Pepsi, Red Bull Zero, and Monster Zero.*

**Part 5: Knowledge, attitudes, and behaviors regarding sugar-sweetened beverages (SSB**

|  |  | None | 1-3 times/per week | 4-6 times/per week | 1 time/per day | 2 times/per day | 3 or more times per day |
| --- | --- | --- | --- | --- | --- | --- | --- |
| (1) | **Sugar-sweetened carbonated beverages with sugar (regular type, not sugar-free or zero-sugar type)**  *e.g. Coca-Cola, Pepsi, Fanta, 7up, 100 plus (carbonated), etc.* |  |  |  |  |  |  |
| (2) | **Sugar-sweetened non-carbonated drinks**  *e.g.* *lemon water, flavored water, coconut water, vitamin water, barley water, etc.* |  |  |  |  |  |  |
| (3) | **Sugar-sweetened energy drinks (regular type, not sugar-free or zero-sugar type)**  *e.g. Red Bull, V, Monster, etc.* |  |  |  |  |  |  |
| (4) | **Sugar-sweetened sports/isotonic drinks**  *e.g. 100 Plus -Active (non-carbonated), Pocari Sweat, Gatorade, etc.* |  |  |  |  |  |  |
| (5) | **Sweetened fruit drinks**  *e.g. apple juice, mango juice, watermelon juice, etc.* |  |  |  |  |  |  |
| (6) | **Sugar-sweetened malted drinks**  *e.g.* *Milo, Ovaltine, Horlicks, etc.* |  |  |  |  |  |  |
| (7) | **Sugar-sweetened ready-to-drink (freshly prepared) tea**  *e.g. Teh,Teh Tarik, lemon tea, milk tea, bubble tea,*  *teas and other drinks with pearls/bubbles/jelly, etc.* |  |  |  |  |  |  |
| (8) | **Sugar-sweetened ready-to-drink (freshly prepared) coffee**  *e.g. Kopi, Kopi O, Kopi C, Yuan Yang,* *coffee frappuccino, etc.* |  |  |  |  |  |  |
| (9) | **Sugar-sweetened canned coffee or tea drinks (with or without milk)**  *e.g. Nescafe, Pokka, Boss, etc.* |  |  |  |  |  |  |
| (10) | **Sugar-sweetened milk drinks**  *e.g. soya milk with sugar, chocolate milk, yogurt drinks, Yakult cultured milk bottle drinks, flavored milk, etc.* |  |  |  |  |  |  |
| (11) | **Traditional Chinese herbal sweetened beverages**  *e.g.*  *Red Date Tea, Ginger tea, Bird's Nest Beverage, etc.* |  |  |  |  |  |  |

20. **In a typical week**, how often do you consume the following sugar-sweetened beverages?

|  |  | None | 1-3 times/per week | 4-6 times/ per week | 1 time/per day | 2 times/per day | 3 or more times per day |
| --- | --- | --- | --- | --- | --- | --- | --- |
| (1) | Artificially sweetened carbonated beverages  e.g. *Coke Zero, 100 Plus Zero, Sprite Zero, Diet Coca-Cola, Diet Pepsi, etc.* |  |  |  |  |  |  |
| (2) | Artificial-sweetened non-carbonated drink  *e.g. Ice lemon tea (Zero Sugar), Ice Peach Tea (Zero Sugar), etc.* |  |  |  |  |  |  |
| (3) | Artificial-sweetened energy drinks  *e.g. Red Bull Zero, V, Monster Zero, etc.* |  |  |  |  |  |  |

21. **In a typical week**, how often do you consume the following artificially sweetened beverages?
