## Appendix B of the Supplementary Material for "Influences of Nutri-Grade front-of-pack labels on the consumption of sugar-sweetened and artificially sweetened beverages: moderating roles of the food environment and age"

**Table S1.** Moderation effects of knowledge of Nutri-Grade labels and built environment factors on SSB consumption daily in Singapore (n = 2,868)

| **Characteristics** | Model 1 | | Model 2 | | Model 3 | | Model 4 | | Model 5 | | Model 6 | | Model 7 | |
| --- | --- | --- | --- | --- | --- | --- | --- | --- | --- | --- | --- | --- | --- | --- |
|  | AOR | 95% CI | AOR | 95% CI | AOR | 95% CI | AOR | 95% CI | AOR | 95% CI | AOR | 95% CI | AOR | 95% CI |
| **Demographic** |  |  |  |  |  |  |  |  |  |  |  |  |  |  |
| **Age** |  |  |  |  |  |  |  |  |  |  |  |  |  |  |
| 21-39 years | Ref |  | Ref |  | Ref |  | Ref |  | Ref |  | Ref |  | Ref |  |
| 40-59 years | 0.82* | 0.69–0.99 | 0.81* | 0.68–0.97 | 0.80* | 0.67–0.96 | 0.81* | 0.68–0.97 | 0.81* | 0.68–0.97 | 0.81* | 0.68–0.97 | 0.80* | 0.67–0.96 |
| 60 years or above | 1.48* | 1.10–1.99 | 1.46* | 1.08–1.97 | 1.43* | 1.06–1.92 | 1.44* | 1.07–1.94 | 1.45* | 1.07–1.95 | 1.47* | 1.09–1.98 | 1.43* | 1.06–1.93 |
| **Sex** |  |  |  |  |  |  |  |  |  |  |  |  |  |  |
| Male | Ref |  | Ref |  | Ref |  | Ref |  | Ref |  | Ref |  | Ref |  |
| Female | 0.78*** | 0.67–0.90 | 0.78*** | 0.68–0.90 | 0.78*** | 0.67–0.90 | 0.78*** | 0.67–0.90 | 0.77*** | 0.66–0.89 | 0.78*** | 0.68–0.91 | 0.78*** | 0.68–0.90 |
| **Ethnicity** |  |  |  |  |  |  |  |  |  |  |  |  |  |  |
| Chinese | Ref |  | Ref |  | Ref |  | Ref |  | Ref |  | Ref |  | Ref |  |
| Malay | 0.72** | 0.59–0.89 | 0.75** | 0.61–0.92 | 0.75** | 0.62–0.93 | 0.75** | 0.61–0.92 | 0.76** | 0.62–0.93 | 0.75** | 0.61–0.92 | 0.76** | 0.62–0.93 |
| Indian | 0.97 | 0.78–1.22 | 0.96 | 0.77–1.20 | 0.96 | 0.77–1.20 | 0.97 | 0.78–1.22 | 0.97 | 0.78–1.22 | 0.96 | 0.77–1.21 | 0.97 | 0.77–1.21 |
| Others | 2.62** | 1.30–5.27 | 2.63** | 1.31–5.29 | 2.62** | 1.30–5.25 | 2.59** | 1.29–5.19 | 2.61** | 1.30–5.23 | 2.56** | 1.28–5.14 | 2.59** | 1.29–5.20 |
| **Education level** |  |  |  |  |  |  |  |  |  |  |  |  |  |  |
| Below university | Ref |  | Ref |  | Ref |  | Ref |  | Ref |  | Ref |  | Ref |  |
| University and above | 0.97 | 0.83–1.13 | 0.96 | 0.82–1.12 | 0.96 | 0.83–1.13 | 0.97 | 0.83–1.13 | 0.96 | 0.82–1.12 | 0.96 | 0.82–1.12 | 0.97 | 0.83–1.13 |
| **Marital status** |  |  |  |  |  |  |  |  |  |  |  |  |  |  |
| Married with children | Ref |  | Ref |  | Ref |  | Ref |  | Ref |  | Ref |  | Ref |  |
| Married without children | 0.95 | 0.74–1.21 | 0.94 | 0.74–1.20 | 0.93 | 0.73–1.19 | 0.93 | 0.73–1.19 | 0.96 | 0.75–1.22 | 0.94 | 0.74–1.20 | 0.93 | 0.73–1.19 |
| Single | 0.85 | 0.70–1.03 | 0.84 | 0.69–1.02 | 0.83 | 0.68–1.01 | 0.83 | 0.68–1.01 | 0.84 | 0.69–1.02 | 0.83 | 0.69–1.01 | 0.84 | 0.69–1.02 |
| Widowed/Divorced/Separated | 0.46*** | 0.33–0.63 | 0.45*** | 0.33–0.62 | 0.44*** | 0.32–0.61 | 0.44*** | 0.32–0.61 | 0.45*** | 0.33–0.62 | 0.45*** | 0.32–0.62 | 0.44*** | 0.32–0.61 |
| **Employment status** |  |  |  |  |  |  |  |  |  |  |  |  |  |  |
| Employed full-time | Ref |  | Ref |  | Ref |  | Ref |  | Ref |  | Ref |  | Ref |  |
| Employed part-time | 1.09 | 0.81–1.46 | 1.09 | 0.81–1.47 | 1.09 | 0.81–1.47 | 1.11 | 0.83–1.50 | 1.09 | 0.81–1.47 | 1.10 | 0.81–1.47 | 1.10 | 0.82–1.48 |
| Others | 0.70 | 0.42–1.19 | 0.73 | 0.43–1.23 | 0.73 | 0.43–1.24 | 0.74 | 0.44–1.25 | 0.74 | 0.44–1.25 | 0.74 | 0.44–1.25 | 0.73 | 0.43–1.24 |
| Retired/Homemaker | 1.09 | 0.84–1.41 | 1.07 | 0.83–1.38 | 1.08 | 0.83–1.39 | 1.09 | 0.84–1.41 | 1.08 | 0.83–1.40 | 1.07 | 0.83–1.38 | 1.08 | 0.83–1.39 |
| Self-employed | 0.59** | 0.42–0.83 | 0.63** | 0.45–0.89 | 0.63** | 0.45–0.89 | 0.63** | 0.45–0.89 | 0.63** | 0.45–0.88 | 0.63** | 0.45–0.89 | 0.64** | 0.45–0.89 |
| Unemployed | 1.35 | 0.85–2.14 | 1.39 | 0.87–2.21 | 1.38 | 0.87–2.19 | 1.40 | 0.88–2.22 | 1.39 | 0.88–2.20 | 1.38 | 0.87–2.20 | 1.36 | 0.86–2.15 |
| **Health status** |  |  |  |  |  |  |  |  |  |  |  |  |  |  |
| **BMI** |  |  |  |  |  |  |  |  |  |  |  |  |  |  |
| Normal 18.5-22.9 | Ref |  | Ref |  | Ref |  | Ref |  | Ref |  | Ref |  | Ref |  |
| Underweight <18.5 | 0.95 | 0.71–1.28 | 0.96 | 0.71–1.29 | 0.95 | 0.71–1.28 | 0.95 | 0.71–1.28 | 0.97 | 0.72–1.30 | 0.96 | 0.71–1.29 | 0.96 | 0.71–1.29 |
| Overweight 23-27.4 | 0.81* | 0.69–0.95 | 0.81* | 0.69–0.95 | 0.81* | 0.69–0.96 | 0.81* | 0.69–0.96 | 0.81* | 0.69–0.96 | 0.82* | 0.69–0.96 | 0.82* | 0.69–0.96 |
| Obese ≥27.5 | 1.13 | 0.88–1.44 | 1.14 | 0.89–1.45 | 1.14 | 0.90–1.46 | 1.13 | 0.89–1.44 | 1.14 | 0.89–1.45 | 1.13 | 0.89–1.45 | 1.14 | 0.89–1.45 |
| **Medical conditions** |  |  |  |  |  |  |  |  |  |  |  |  |  |  |
| Yes | Ref |  | Ref |  | Ref |  | Ref |  | Ref |  | Ref |  | Ref |  |
| No | 1.27** | 1.08–1.50 | 1.27** | 1.08–1.50 | 1.29** | 1.10–1.52 | 1.27** | 1.08–1.49 | 1.26** | 1.07–1.49 | 1.27** | 1.08–1.49 | 1.29** | 1.10–1.52 |
| **Individuals’ knowledge and belief** |  |  |  |  |  |  |  |  |  |  |  |  |  |  |
| Knowledge of SSBs | 0.84*** | 0.78–0.92 | 0.84*** | 0.77–0.91 | 0.84*** | 0.77–0.91 | 0.84*** | 0.77–0.91 | 0.85*** | 0.78–0.92 | 0.84*** | 0.77–0.91 | 0.84*** | 0.77–0.91 |
| Perceived control of SSB intake | 0.80*** | 0.73–0.89 | 0.81*** | 0.73–0.90 | 0.81*** | 0.74–0.90 | 0.81*** | 0.73–0.89 | 0.81*** | 0.73–0.89 | 0.81*** | 0.73–0.90 | 0.81*** | 0.73–0.90 |
| Self-efficacy | 0.95 | 0.86–1.05 | 0.96 | 0.87–1.06 | 0.97 | 0.88–1.07 | 0.96 | 0.87–1.06 | 0.96 | 0.87–1.05 | 0.96 | 0.87–1.06 | 0.97 | 0.88–1.07 |
| **Physical environment** |  |  |  |  |  |  |  |  |  |  |  |  |  |  |
| Household availability of SSBs |  |  |  |  |  |  |  |  |  |  |  |  |  |  |
| Very rarely | Ref |  | Ref |  | Ref |  | Ref |  | Ref |  | Ref |  | Ref |  |
| Monthly | 3.04*** | 2.31–4.02 | 3.06*** | 2.32–4.04 | 3.09*** | 2.34–4.08 | 3.07*** | 2.33–4.06 | 3.01*** | 2.28–3.98 | 3.08*** | 2.34–4.07 | 3.09*** | 2.34–4.08 |
| Weekly | 3.62*** | 2.70–4.85 | 3.69*** | 2.75–4.94 | 3.67*** | 2.74–4.92 | 3.79*** | 2.83–5.08 | 3.66*** | 2.74–4.91 | 3.76*** | 2.81–5.05 | 3.66*** | 2.73–4.91 |
| Every other day/daily | 4.40*** | 3.30–5.85 | 4.45*** | 3.34–5.92 | 4.41*** | 3.32–5.88 | 4.49*** | 3.37–5.98 | 4.43*** | 3.33–5.90 | 4.50*** | 3.38–5.99 | 4.42*** | 3.32–5.88 |
| Purchase of SSBs via online delivery |  |  |  |  |  |  |  |  |  |  |  |  |  |  |
| Very rarely | Ref |  | Ref |  | Ref |  | Ref |  | Ref |  | Ref |  | Ref |  |
| Monthly | 2.56*** | 2.03–3.23 | 2.54*** | 2.02–3.20 | 2.59*** | 2.06–3.27 | 2.52*** | 2.00–3.18 | 2.60*** | 2.06–3.27 | 2.53*** | 2.01–3.19 | 2.57*** | 2.04–3.24 |
| Weekly | 2.89*** | 2.27–3.69 | 2.91*** | 2.28–3.71 | 2.93*** | 2.29–3.73 | 2.85*** | 2.24–3.64 | 2.93*** | 2.30–3.74 | 2.88*** | 2.26–3.68 | 2.92*** | 2.29–3.72 |
| Every other day/daily | 2.40*** | 1.92–3.02 | 2.39*** | 1.91–3.00 | 2.42*** | 1.93–3.03 | 2.37*** | 1.88–2.97 | 2.43*** | 1.94–3.05 | 2.38*** | 1.90–2.99 | 2.42*** | 1.93–3.04 |
| **Social environment** |  |  |  |  |  |  |  |  |  |  |  |  |  |  |
| Perceived social norm of acceptance of SSB intake | 1.54*** | 1.41–1.69 | 1.53*** | 1.40–1.68 | 1.52*** | 1.39–1.67 | 1.53*** | 1.40–1.68 | 1.52*** | 1.39–1.67 | 1.53*** | 1.39–1.67 | 1.52*** | 1.39–1.67 |
| Exposure to SSB advertising | 1.24*** | 1.13–1.36 | 1.23*** | 1.12–1.35 | 1.24*** | 1.13–1.36 | 1.24*** | 1.13–1.36 | 1.24*** | 1.13–1.36 | 1.24*** | 1.13–1.36 | 1.24*** | 1.13–1.36 |
| **Nutri-Grade FOPL influences** |  |  |  |  |  |  |  |  |  |  |  |  |  |  |
| Perception of Nutri-Grade labels | 0.73*** | 0.67–0.78 | 0.72*** | 0.67–0.78 | 0.72*** | 0.67–0.78 | 0.72*** | 0.67–0.78 | 0.72*** | 0.67–0.78 | 0.72*** | 0.67–0.78 | 0.72*** | 0.67–0.78 |
| Knowledge of the Nutri-Grade labels | 1.03 | 0.95–1.12 | 1.04 | 0.96–1.13 | 1.04 | 0.96–1.13 | 1.05 | 0.96–1.14 | 1.03 | 0.95–1.12 | 1.04 | 0.95–1.13 | 1.04 | 0.96–1.14 |
| **Built environment** |  |  |  |  |  |  |  |  |  |  |  |  |  |  |
| HDB house price | 0.89 | 0.77–1.02 | 0.85* | 0.74–0.98 | 0.84* | 0.73–0.97 | 0**.**83* | 0.71–0.96 | 0.83** | 0.72–0.95 | 0.86* | 0.74–1.00 | 0.83* | 0.70–0.97 |
| Commercial land use | 1.17* | 1.03–1.33 | 1.15* | 1.01–1.31 | 1.13 | 0.99–1.28 | 1.14 | 0.99–1.30 | 1.12 | 0.97–1.30 | 1.18* | 1.01–1.39 | 1.14 | 0.98–1.32 |
| Density of Food Courts (per km^2^) | 1.06 | 0.94–1.21 | 1.10 | 0.96–1.24 | 1.09 | 0.95–1.24 |  |  |  |  |  |  |  |  |
| Density of Bars (per km^2^) |  |  |  |  |  |  | 0.98 | 0.89–1.09 |  |  |  |  |  |  |
| Density of Cafe (per km^2^) |  |  |  |  |  |  |  |  | 1.05 | 0.91–1.20 |  |  |  |  |
| Density of vending machines (per km^2^) |  |  |  |  |  |  |  |  |  |  | 0.93 | 0.80–1.09 |  |  |
| Density of juice bars (per km^2^) |  |  |  |  |  |  |  |  |  |  |  |  | 0.96 | 0.83–1.10 |
| **Interaction** |  |  |  |  |  |  |  |  |  |  |  |  |  |  |
| Knowledge of Nutri-Grade labels×HDB house price | 1.30*** | 1.21–1.41 |  |  |  |  |  |  |  |  |  |  |  |  |
| Knowledge of Nutri-Grade labels×commercial land use |  |  | 1.09* | 1.02–1.18 |  |  |  |  |  |  |  |  |  |  |
| Knowledge of Nutri-Grade labels×food courts |  |  |  |  | 1.00 | 0.93–1.07 |  |  |  |  |  |  |  |  |
| Knowledge of Nutri-Grade labels×bars |  |  |  |  |  |  | 1.17*** | 1.07–1.27 |  |  |  |  |  |  |
| Knowledge of Nutri-Grade labels×cafes |  |  |  |  |  |  |  |  | 1.13** | 1.05–1.22 |  |  |  |  |
| Knowledge of Nutri-Grade labels×vending machines |  |  |  |  |  |  |  |  |  |  | 1.09* | 1.01–1.18 |  |  |
| Knowledge of Nutri-Grade labels×juice bars |  |  |  |  |  |  |  |  |  |  |  |  | 0.96 | 0.89–1.04 |
| AIC | 6898.58 |  | 6941.13 |  | 6946.74 |  | 6935.55 |  | 6938.19 |  | 6941.98 |  | 6946.76 |  |
| Log-Likelihood | -3408.29 |  | -3429.56 |  | -3432.37 |  | -3426.77 |  | -3428.10 |  | -3429.99 |  | -3432.38 |  |

AOR: Adjusted Odds Ratio; CI: Confidence Interval; **p* < .05; ***p* < .01; ****p* < .001.

**Table S2.** Moderation effects of perception of Nutri-Grade labels and built environment factors on daily sugar-sweetened beverage consumption in Singapore (n = 2,868)

| **Characteristics** | Model 1 | | Model 2 | | Model 3 | | | Model 4 | | | Model 5 | | | Model 6 | | Model 7 | |
| --- | --- | --- | --- | --- | --- | --- | --- | --- | --- | --- | --- | --- | --- | --- | --- | --- | --- |
|  | AOR | 95% CI | AOR | 95% CI | AOR | 95% CI | AOR | | 95% CI | AOR | | 95% CI | AOR | | 95% CI | AOR | 95% CI |
| **Demographic** |  |  |  |  |  |  |  | |  |  | |  |  | |  |  |  |
| **Age** |  |  |  |  |  |  |  | |  |  | |  |  | |  |  |  |
| 21-39 years | Ref |  | Ref |  | Ref |  | Ref | |  | Ref | |  | Ref | |  | Ref |  |
| 40-59 years | 0.80* | 0.67–0.96 | 0.80* | 0.67–0.96 | 0.80* | 0.67–0.96 | 0.80* | | 0.67–0.96 | 0.80* | | 0.67–0.96 | 0.80* | | 0.67–0.96 | 0.80* | 0.67–0.96 |
| 60 years or above | 1.43* | 1.06–1.92 | 1.43* | 1.06–1.92 | 1.43* | 1.06–1.93 | 1.42* | | 1.06–1.92 | 1.43* | | 1.06–1.92 | 1.43* | | 1.06–1.93 | 1.43* | 1.06–1.92 |
| **Sex** |  |  |  |  |  |  |  | |  |  | |  |  | |  |  |  |
| Male | Ref |  | Ref |  | Ref |  | Ref | |  | Ref | |  | Ref | |  | Ref |  |
| Female | 0.78*** | 0.67–0.90 | 0.78*** | 0.67–0.90 | 0.79** | 0.68–0.91 | 0.78*** | | 0.67–0.90 | 0.78*** | | 0.68–0.90 | 0.78*** | | 0.68–0.91 | 0.78** | 0.68–0.91 |
| **Ethnicity** |  |  |  |  |  |  |  | |  |  | |  |  | |  |  |  |
| Chinese | Ref |  | Ref |  | Ref |  | Ref | |  | Ref | |  | Ref | |  | Ref |  |
| Malay | 0.75** | 0.62–0.93 | 0.75** | 0.62–0.93 | 0.76** | 0.62–0.93 | 0.75** | | 0.62–0.93 | 0.76** | | 0.62–0.93 | 0.76** | | 0.62–0.93 | 0.76** | 0.62–0.93 |
| Indian | 0.96 | 0.77–1.20 | 0.96 | 0.76–1.20 | 0.95 | 0.76–1.19 | 0.96 | | 0.77–1.20 | 0.96 | | 0.77–1.21 | 0.97 | | 0.77–1.21 | 0.96 | 0.77–1.21 |
| Others | 2.62** | 1.30–5.25 | 2.61** | 1.30–5.23 | 2.50** | 1.25–5.01 | 2.59** | | 1.29–5.20 | 2.60** | | 1.30–5.23 | 2.58** | | 1.29–5.18 | 2.57** | 1.28–5.16 |
| **Education level** |  |  |  |  |  |  |  | |  |  | |  |  | |  |  |  |
| Below university | Ref |  | Ref |  | Ref |  | Ref | |  | Ref | |  | Ref | |  | Ref |  |
| University and above | 0.96 | 0.83–1.13 | 0.96 | 0.83–1.13 | 0.97 | 0.83–1.14 | 0.96 | | 0.82–1.12 | 0.96 | | 0.83–1.13 | 0.97 | | 0.83–1.13 | 0.96 | 0.83–1.13 |
| **Marital status** |  |  |  |  |  |  |  | |  |  | |  |  | |  |  |  |
| Married with children | Ref |  | Ref |  | Ref |  | Ref | |  | Ref | |  | Ref | |  | Ref |  |
| Married without children | 0.93 | 0.73–1.19 | 0.93 | 0.73–1.18 | 0.93 | 0.73–1.18 | 0.93 | | 0.73–1.18 | 0.93 | | 0.73–1.19 | 0.93 | | 0.73–1.19 | 0.93 | 0.73–1.19 |
| Single | 0.83 | 0.68–1.01 | 0.83 | 0.68–1.01 | 0.83 | 0.68–1.01 | 0.83 | | 0.68–1.01 | 0.84 | | 0.69–1.02 | 0.84 | | 0.69–1.02 | 0.84 | 0.69–1.02 |
| Widowed/Divorced/Separated | 0.44*** | 0.32–0.61 | 0.44*** | 0.32–0.61 | 0.44*** | 0.32–0.61 | 0.44*** | | 0.32–0.61 | 0.44*** | | 0.32–0.61 | 0.44*** | | 0.32–0.61 | 0.44*** | 0.32–0.61 |
| **Employment status** |  |  |  |  |  |  |  | |  |  | |  |  | |  |  |  |
| Employed full-time | Ref |  | Ref |  | Ref |  | Ref | |  | Ref | |  | Ref | |  | Ref |  |
| Employed part-time | 1.09 | 0.81–1.47 | 1.09 | 0.81–1.47 | 1.09 | 0.81–1.47 | 1.09 | | 0.81–1.47 | 1.10 | | 0.82–1.48 | 1.10 | | 0.81–1.47 | 1.10 | 0.82–1.48 |
| Others | 0.73 | 0.43–1.24 | 0.73 | 0.43–1.24 | 0.74 | 0.44–1.25 | 0.73 | | 0.43–1.24 | 0.73 | | 0.43–1.24 | 0.73 | | 0.43–1.23 | 0.74 | 0.43–1.24 |
| Retired/Homemaker | 1.08 | 0.83–1.39 | 1.07 | 0.83–1.39 | 1.05 | 0.81–1.35 | 1.07 | | 0.83–1.39 | 1.07 | | 0.83–1.39 | 1.07 | | 0.83–1.38 | 1.07 | 0.83–1.38 |
| Self-employed | 0.63** | 0.45–0.89 | 0.63** | 0.45–0.88 | 0.61** | 0.44–0.86 | 0.64** | | 0.45–0.90 | 0.63** | | 0.45–0.88 | 0.63** | | 0.45–0.89 | 0.63** | 0.45–0.89 |
| Unemployed | 1.38 | 0.87–2.19 | 1.37 | 0.87–2.18 | 1.38 | 0.87–2.19 | 1.36 | | 0.86–2.16 | 1.39 | | 0.87–2.20 | 1.37 | | 0.87–2.18 | 1.37 | 0.86–2.17 |
| **Health status** |  |  |  |  |  |  |  | |  |  | |  |  | |  |  |  |
| **BMI** |  |  |  |  |  |  |  | |  |  | |  |  | |  |  |  |
| Normal 18.5-22.9 | Ref |  | Ref |  | Ref |  | Ref | |  | Ref | |  | Ref | |  | Ref |  |
| Underweight <18.5 | 0.96 | 0.71–1.28 | 0.95 | 0.71–1.28 | 0.97 | 0.72–1.31 | 0.96 | | 0.71–1.28 | 0.96 | | 0.72–1.29 | 0.96 | | 0.71–1.29 | 0.96 | 0.72–1.29 |
| Overweight 23-27.4 | 0.81* | 0.69–0.96 | 0.81* | 0.69–0.96 | 0.82* | 0.70–0.97 | 0.81* | | 0.69–0.96 | 0.81* | | 0.69–0.96 | 0.82* | | 0.69–0.96 | 0.82* | 0.69–0.96 |
| Obese ≥27.5 | 1.14 | 0.90–1.46 | 1.15 | 0.90–1.46 | 1.17 | 0.91–1.49 | 1.14 | | 0.89–1.45 | 1.14 | | 0.90–1.46 | 1.14 | | 0.89–1.46 | 1.14 | 0.90–1.46 |
| **Medical conditions** |  |  |  |  |  |  |  | |  |  | |  |  | |  |  |  |
| Yes | Ref |  | Ref |  | Ref |  | Ref | |  | Ref | |  | Ref | |  | Ref |  |
| No | 1.29** | 1.10–1.52 | 1.29** | 1.09–1.52 | 1.30** | 1.10–1.53 | 1.29** | | 1.09–1.52 | 1.29** | | 1.10–1.52 | 1.29** | | 1.10–1.52 | 1.29** | 1.10–1.52 |
| **Individuals’ knowledge and belief** |  |  |  |  |  |  |  | |  |  | |  |  | |  |  |  |
| Knowledge of SSBs | 0.84*** | 0.77–0.91 | 0.84*** | 0.77–0.91 | 0.85*** | 0.78–0.92 | 0.84*** | | 0.77–0.91 | 0.84*** | | 0.77–0.91 | 0.84*** | | 0.77–0.91 | 0.84*** | 0.77–0.91 |
| Perceived control of SSB intake | 0.81*** | 0.74–0.90 | 0.81*** | 0.74–0.90 | 0.81*** | 0.74–0.90 | 0.81*** | | 0.74–0.90 | 0.81*** | | 0.73–0.90 | 0.81*** | | 0.74–0.90 | 0.81*** | 0.73–0.90 |
| Self-efficacy | 0.97 | 0.88–1.07 | 0.97 | 0.88–1.07 | 0.96 | 0.87–1.06 | 0.97 | | 0.88–1.07 | 0.97 | | 0.88–1.07 | 0.97 | | 0.88–1.07 | 0.97 | 0.88–1.07 |
| **Physical environment** |  |  |  |  |  |  |  | |  |  | |  |  | |  |  |  |
| Household availability of SSBs |  |  |  |  |  |  |  | |  |  | |  |  | |  |  |  |
| Very rarely | Ref |  | Ref |  | Ref |  | Ref | |  | Ref | |  | Ref | |  | Ref |  |
| Monthly | 3.09*** | 2.34–4.08 | 3.10*** | 2.35–4.09 | 3.11*** | 2.35–4.10 | 3.10*** | | 2.35–4.10 | 3.06*** | | 2.32–4.04 | 3.08*** | | 2.33–4.07 | 3.08*** | 2.33–4.06 |
| Weekly | 3.67*** | 2.74–4.92 | 3.68*** | 2.75–4.93 | 3.73*** | 2.78–5.00 | 3.72*** | | 2.77–4.98 | 3.66*** | | 2.73–4.90 | 3.71*** | | 2.77–4.97 | 3.68*** | 2.75–4.93 |
| Every other day/daily | 4.41*** | 3.31–5.87 | 4.42*** | 3.32–5.88 | 4.41*** | 3.31–5.87 | 4.45*** | | 3.34–5.93 | 4.41*** | | 3.31–5.87 | 4.45*** | | 3.34–5.93 | 4.42*** | 3.32–5.89 |
| Purchase of SSBs via online delivery |  |  |  |  |  |  |  | |  |  | |  |  | |  |  |  |
| Very rarely | Ref |  | Ref |  | Ref |  | Ref | |  | Ref | |  | Ref | |  | Ref |  |
| Monthly | 2.59*** | 2.06–3.27 | 2.59*** | 2.06–3.27 | 2.60*** | 2.07–3.28 | 2.56*** | | 2.03–3.22 | 2.58*** | | 2.05–3.25 | 2.55*** | | 2.02–3.22 | 2.57*** | 2.04–3.23 |
| Weekly | 2.93*** | 2.29–3.73 | 2.94*** | 2.30–3.75 | 2.97*** | 2.33–3.79 | 2.89*** | | 2.27–3.69 | 2.92*** | | 2.29–3.72 | 2.89*** | | 2.27–3.69 | 2.91*** | 2.28–3.72 |
| Every other day/daily | 2.42*** | 1.93–3.03 | 2.42*** | 1.93–3.04 | 2.39*** | 1.91–3.00 | 2.41*** | | 1.92–3.02 | 2.41*** | | 1.92–3.03 | 2.40*** | | 1.91–3.02 | 2.41*** | 1.92–3.03 |
| **Social environment** |  |  |  |  |  |  |  | |  |  | |  |  | |  |  |  |
| Perceived social norm of acceptance of SSB intake | 1.52*** | 1.39–1.67 | 1.52*** | 1.39–1.67 | 1.52*** | 1.38–1.66 | 1.52*** | | 1.39–1.67 | 1.52*** | | 1.39–1.67 | 1.52*** | | 1.39–1.67 | 1.52*** | 1.39–1.66 |
| Exposure to SSB advertising | 1.24*** | 1.13–1.36 | 1.23*** | 1.13–1.35 | 1.23*** | 1.12–1.35 | 1.24*** | | 1.13–1.36 | 1.24*** | | 1.13–1.36 | 1.24*** | | 1.13–1.36 | 1.24*** | 1.13–1.36 |
| **Nutri-Grade FOPL influences** |  |  |  |  |  |  |  | |  |  | |  |  | |  |  |  |
| Perception of Nutri-Grade labels | 0.72*** | 0.67–0.78 | 0.72*** | 0.67–0.78 | 0.72*** | 0.66–0.77 | 0.72*** | | 0.67–0.78 | 0.72*** | | 0.67–0.78 | 0.73*** | | 0.67–0.78 | 0.72*** | 0.67–0.78 |
| Knowledge of the Nutri-Grade labels | 1.04 | 0.96–1.13 | 1.04 | 0.96–1.13 | 1.03 | 0.95–1.12 | 1.04 | | 0.95–1.13 | 1.04 | | 0.96–1.13 | 1.03 | | 0.95–1.12 | 1.04 | 0.95–1.13 |
| **Built environment** |  |  |  |  |  |  |  | |  |  | |  |  | |  |  |  |
| HDB house price | 0.84* | 0.73–0.97 | 0.84* | 0.73–0.97 | 0.85* | 0.74–0.98 | 0.82** | | 0.71–0.95 | 0.84* | | 0.72–0.97 | 0.85* | | 0.73–0.99 | 0.82* | 0.70–0.97 |
| Commercial land use | 1.13 | 0.99–1.28 | 1.12 | 0.98–1.28 | 1.11 | 0.98–1.27 | 1.13 | | 0.99–1.30 | 1.10 | | 0.95–1.29 | 1.18* | | 1.01–1.39 | 1.14 | 0.98–1.33 |
| Density of Food Courts (per km^2^) | 1.09 | 0.95–1.24 | 1.09 | 0.95–1.24 | 1.10 | 0.96–1.25 |  | |  |  | |  |  | |  |  |  |
| Density of Bars (per km^2^) |  |  |  |  |  |  | 0.96 | | 0.86–1.06 |  | |  |  | |  |  |  |
| Density of Cafe (per km^2^) |  |  |  |  |  |  |  | |  | 1.03 | | 0.89–1.18 |  | |  |  |  |
| Density of vending machines (per km^2^) |  |  |  |  |  |  |  | |  |  | |  | 0.91 | | 0.78–1.07 |  |  |
| Density of juice bars (per km^2^) |  |  |  |  |  |  |  | |  |  | |  |  | |  | 0.96 | 0.83–1.11 |
| **Interaction** |  |  |  |  |  |  |  | |  |  | |  |  | |  |  |  |
| Perception of Nutri-Grade labels×HDB house price | 1.00 | 0.93–1.08 |  |  |  |  |  | |  |  | |  |  | |  |  |  |
| Perception of Nutri-Grade labels×Commercial land use |  |  | 0.98 | 0.90–1.05 |  |  |  | |  |  | |  |  | |  |  |  |
| Perception of Nutri-Grade labels×Food Courts |  |  |  |  | 1.12** | 1.04–1.21 |  | |  |  | |  |  | |  |  |  |
| Perception of Nutri-Grade labels×Bars |  |  |  |  |  |  | 0.97 | | 0.89–1.06 |  | |  |  | |  |  |  |
| Perception of Nutri-Grade labels×Cafes |  |  |  |  |  |  |  | |  | 1.03 | | 0.96–1.11 |  | |  |  |  |
| Perception of Nutri-Grade labels×Vending machines |  |  |  |  |  |  |  | |  |  | |  | 1.02 | | 0.94–1.10 |  |  |
| Perception of Nutri-Grade labels×juice bars |  |  |  |  |  |  |  | |  |  | |  |  | |  | 1.03 | 0.96–1.11 |
| AIC | 6946.74 |  | 6946.35 |  | 6938.23 |  | 6946.69 | |  | 6947.41 | |  | 6946.70 | |  | 6947.11 |  |
| Log-Likelihood | -3432.37 |  | -3432.17 |  | -3428.11 |  | -3432.35 | |  | -3432.70 | |  | -3432.35 | |  | -3432.55 |  |

AOR: Adjusted Odds Ratio; CI: Confidence Interval; **p* < .05; ***p* < .01; ****p* < .001.

**Table S3.** Moderation effects of knowledge of Nutri-Grade labels and built environment factors on the total number of different types of SSBs consumed weekly in Singapore (n = 2,868)

| **Characteristics** | Model 1 | | Model 2 | | Model 3 | | | Model 4 | | Model 5 | | | Model 6 | | | Model 7 | |
| --- | --- | --- | --- | --- | --- | --- | --- | --- | --- | --- | --- | --- | --- | --- | --- | --- | --- |
|  | IRR | 95% CI | IRR | 95% CI | IRR | 95% CI | | IRR | 95% CI | IRR | 95% CI | IRR | | 95% CI | IRR | | 95% CI |
| **Demographic** |  |  |  |  |  |  | |  |  |  |  |  | |  |  | |  |
| **Age** |  |  |  |  |  |  | |  |  |  |  |  | |  |  | |  |
| 21-39 years | Ref |  | Ref |  | Ref |  | | Ref |  | Ref |  | Ref | |  | Ref | |  |
| 40-59 years | 0.92*** | 0.88–0.96 | 0.91*** | 0.87–0.95 | 0.91*** | 0.87–0.95 | | 0.91*** | 0.87–0.95 | 0.91*** | 0.87–0.95 | 0.91*** | | 0.87–0.95 | 0.91*** | | 0.87–0.95 |
| 60 years or above | 0.95 | 0.88–1.02 | 0.95 | 0.88–1.02 | 0.94 | | 0.88–1.01 | 0.94 | 0.88–1.01 | 0.94 | 0.88–1.01 | 0.95 | | 0.88–1.02 | 0.95 | | 0.88–1.02 |
| **Sex** |  |  |  |  |  | |  |  |  |  |  |  | |  |  | |  |
| Male | Ref |  | Ref |  | Ref | |  | Ref |  | Ref |  | Ref | |  | Ref | |  |
| Female | 0.94** | 0.91–0.98 | 0.94** | 0.91–0.98 | 0.94** | | 0.91–0.98 | 0.94** | 0.91–0.98 | 0.94*** | 0.91–0.97 | 0.94** | | 0.91–0.98 | 0.94** | | 0.91–0.98 |
| **Ethnicity** |  |  |  |  |  | |  |  |  |  |  |  | |  |  | |  |
| Chinese | Ref |  | Ref |  | Ref | |  | Ref |  | Ref |  | Ref | |  | Ref | |  |
| Malay | 0.89*** | 0.85–0.93 | 0.89*** | 0.85–0.94 | 0.89*** | | 0.85–0.94 | 0.89*** | 0.85–0.94 | 0.89*** | 0.85–0.94 | 0.89*** | | 0.85–0.94 | 0.89*** | | 0.85–0.94 |
| Indian | 0.90*** | 0.85–0.95 | 0.89*** | 0.84–0.94 | 0.89*** | | 0.84–0.94 | 0.89*** | 0.84–0.94 | 0.89*** | 0.84–0.94 | 0.89*** | | 0.84–0.94 | 0.89*** | | 0.84–0.94 |
| Others | 1.11 | 0.95–1.30 | 1.11 | 0.95–1.30 | 1.12 | | 0.96–1.31 | 1.11 | 0.95–1.30 | 1.11 | 0.95–1.30 | 1.11 | | 0.95–1.30 | 1.12 | | 0.95–1.30 |
| **Education level** |  |  |  |  |  | |  |  |  |  |  |  | |  |  | |  |
| Below university | Ref |  | Ref |  | Ref | |  | Ref |  | Ref |  | Ref | |  | Ref | |  |
| University and above | 1.04* | 1.01–1.08 | 1.04* | 1.00–1.08 | 1.04* | | 1.00–1.08 | 1.04* | 1.00–1.08 | 1.04* | 1.00–1.08 | 1.04* | | 1.00–1.08 | 1.04* | | 1.00–1.08 |
| **Marital status** |  |  |  |  |  | |  |  |  |  |  |  | |  |  | |  |
| Married with children | Ref |  | Ref |  | Ref | |  | Ref |  | Ref |  | Ref | |  | Ref | |  |
| Married without children | 1.05 | 1.00–1.12 | 1.05 | 0.99–1.11 | 1.05 | | 0.99–1.11 | 1.05 | 0.99–1.11 | 1.06 | 1.00–1.12 | 1.05 | | 0.99–1.11 | 1.05 | | 0.99–1.11 |
| Single | 0.99 | 0.95–1.04 | 0.99 | 0.95–1.04 | 0.99 | | 0.94–1.04 | 0.99 | 0.94–1.04 | 0.99 | 0.94–1.04 | 0.99 | | 0.94–1.04 | 0.99 | | 0.94–1.04 |
| Widowed/Divorced/Separated | 0.83*** | 0.76–0.90 | 0.82*** | 0.76–0.90 | 0.82*** | | 0.75–0.90 | 0.82*** | 0.75–0.90 | 0.83*** | 0.76–0.90 | 0.82*** | | 0.75–0.90 | 0.82*** | | 0.75–0.89 |
| **Employment status** |  |  |  |  |  | |  |  |  |  |  |  | |  |  | |  |
| Employed full-time | Ref |  | Ref |  | Ref | |  | Ref |  | Ref |  | Ref | |  | Ref | |  |
| Employed part-time | 1.00 | 0.93–1.07 | 1.00 | 0.93–1.08 | 1.00 | | 0.93–1.08 | 1.00 | 0.93–1.08 | 1.00 | 0.93–1.08 | 1.00 | | 0.93–1.08 | 1.00 | | 0.93–1.08 |
| Others | 0.97 | 0.86–1.10 | 0.98 | 0.86–1.11 | 0.98 | | 0.86–1.11 | 0.98 | 0.86–1.11 | 0.98 | 0.86–1.12 | 0.98 | | 0.86–1.11 | 0.98 | | 0.86–1.11 |
| Retired/Homemaker | 1.02 | 0.96–1.09 | 1.02 | 0.96–1.09 | 1.02 | | 0.96–1.09 | 1.02 | 0.96–1.09 | 1.02 | 0.96–1.09 | 1.02 | | 0.96–1.09 | 1.02 | | 0.96–1.09 |
| Self-employed | 0.85*** | 0.78–0.93 | 0.86** | 0.79–0.94 | 0.86** | | 0.78–0.94 | 0.86*** | 0.78–0.94 | 0.86*** | 0.78–0.94 | 0.86** | | 0.79–0.94 | 0.86** | | 0.79–0.94 |
| Unemployed | 1.06 | 0.95–1.19 | 1.07 | 0.95–1.19 | 1.07 | | 0.95–1.19 | 1.07 | 0.96–1.20 | 1.07 | 0.96–1.20 | 1.07 | | 0.95–1.19 | 1.06 | | 0.95–1.19 |
| **Health status** |  |  |  |  |  | |  |  |  |  |  |  | |  |  | |  |
| **BMI** |  |  |  |  |  | |  |  |  |  |  |  | |  |  | |  |
| Normal 18.5-22.9 | Ref |  | Ref |  | Ref | |  | Ref |  | Ref |  | Ref | |  | Ref | |  |
| Underweight <18.5 | 1.02 | 0.95–1.09 | 1.02 | 0.95–1.09 | 1.02 | | 0.95–1.09 | 1.02 | 0.95–1.09 | 1.02 | 0.95–1.10 | 1.02 | | 0.95–1.09 | 1.02 | | 0.95–1.09 |
| Overweight 23-27.4 | 1.00 | 0.96–1.04 | 1.00 | 0.96–1.04 | 1.00 | | 0.96–1.04 | 1.00 | 0.96–1.04 | 1.00 | 0.96–1.04 | 1.00 | | 0.96–1.04 | 1.00 | | 0.96–1.04 |
| Obese ≥27.5 | 1.08* | 1.02–1.14 | 1.08* | 1.02–1.14 | 1.08* | | 1.02–1.14 | 1.08* | 1.02–1.14 | 1.08* | 1.02–1.14 | 1.08* | | 1.02–1.14 | 1.08* | | 1.02–1.14 |
| **Medical conditions** |  |  |  |  |  | |  |  |  |  |  |  | |  |  | |  |
| Yes | Ref |  | Ref |  | Ref | |  | Ref |  | Ref |  | Ref | |  | Ref | |  |
| No | 1.04* | 1.00–1.09 | 1.04* | 1.00–1.09 | 1.04* | | 1.00–1.09 | 1.04* | 1.00–1.08 | 1.04* | 1.00–1.08 | 1.04* | | 1.00–1.09 | 1.05* | | 1.01–1.09 |
| **Individuals’ knowledge and belief** |  |  |  |  |  | |  |  |  |  |  |  | |  |  | |  |
| Knowledge of SSBs | 0.96*** | 0.94–0.98 | 0.96*** | 0.94–0.98 | 0.96*** | | 0.94–0.98 | 0.96*** | 0.94–0.98 | 0.96*** | 0.94–0.98 | 0.96*** | | 0.94–0.98 | 0.96*** | | 0.94–0.97 |
| Perceived control of SSB intake | 0.97* | 0.95–0.99 | 0.97* | 0.95–1.00 | 0.97* | | 0.95–1.00 | 0.97* | 0.95–0.99 | 0.97* | 0.95–1.00 | 0.97* | | 0.95–1.00 | 0.97* | | 0.95–0.99 |
| Self-efficacy | 0.96** | 0.94–0.99 | 0.97** | 0.94–0.99 | 0.97** | | 0.95–0.99 | 0.97** | 0.94–0.99 | 0.97** | 0.94–0.99 | 0.97** | | 0.94–0.99 | 0.97** | | 0.95–0.99 |
| **Physical environment** |  |  |  |  |  | |  |  |  |  |  |  | |  |  | |  |
| Household availability of SSBs |  |  |  |  |  | |  |  |  |  |  |  | |  |  | |  |
| Very rarely | Ref |  | Ref |  | Ref | |  | Ref |  | Ref |  | Ref | |  | Ref | |  |
| Monthly | 1.50*** | 1.39–1.63 | 1.51*** | 1.39–1.63 | 1.51*** | | 1.39–1.63 | 1.51*** | 1.39–1.63 | 1.50*** | 1.39–1.63 | 1.51*** | | 1.39–1.63 | 1.51*** | | 1.40–1.63 |
| Weekly | 1.58*** | 1.46–1.71 | 1.59*** | 1.47–1.72 | 1.58*** | | 1.46–1.71 | 1.59*** | 1.47–1.72 | 1.58*** | 1.46–1.71 | 1.59*** | | 1.47–1.72 | 1.58*** | | 1.46–1.71 |
| Every other day/daily | 1.61*** | 1.49–1.75 | 1.62*** | 1.50–1.75 | 1.61*** | | 1.49–1.74 | 1.62*** | 1.50–1.75 | 1.61*** | 1.49–1.75 | 1.62*** | | 1.50–1.75 | 1.61*** | | 1.49–1.74 |
| Purchase of SSBs via online delivery |  |  |  |  |  | |  |  |  |  |  |  | |  |  | |  |
| Very rarely | Ref |  | Ref |  | Ref | |  | Ref |  | Ref |  | Ref | |  | Ref | |  |
| Monthly | 1.43*** | 1.35–1.51 | 1.43*** | 1.35–1.51 | 1.44*** | | 1.36–1.52 | 1.43*** | 1.35–1.52 | 1.44*** | 1.36–1.52 | 1.43*** | | 1.35–1.51 | 1.44*** | | 1.36–1.52 |
| Weekly | 1.49*** | 1.40–1.58 | 1.49*** | 1.40–1.58 | 1.50*** | | 1.41–1.59 | 1.49*** | 1.40–1.58 | 1.50*** | 1.41–1.59 | 1.49*** | | 1.40–1.58 | 1.50*** | | 1.41–1.59 |
| Every other day/daily | 1.37*** | 1.29–1.45 | 1.37*** | 1.29–1.45 | 1.38*** | | 1.30–1.46 | 1.37*** | 1.29–1.45 | 1.38*** | 1.30–1.46 | 1.37*** | | 1.29–1.45 | 1.38*** | | 1.30–1.46 |
| **Social environment** |  |  |  |  |  | |  |  |  |  |  |  | |  |  | |  |
| Perceived social norm of acceptance of SSB intake | 1.09*** | 1.06–1.11 | 1.09*** | 1.06–1.11 | 1.08*** | | 1.06–1.11 | 1.09*** | 1.06–1.11 | 1.08*** | 1.06–1.11 | 1.09*** | | 1.06–1.11 | 1.08*** | | 1.06–1.11 |
| Exposure to SSB advertising | 1.05*** | 1.03–1.08 | 1.05*** | 1.03–1.08 | 1.05*** | | 1.03–1.08 | 1.05*** | 1.03–1.08 | 1.05*** | 1.03–1.08 | 1.05*** | | 1.03–1.08 | 1.06*** | | 1.03–1.08 |
| **Nutri-Grade FOPL influences** |  |  |  |  |  | |  |  |  |  |  |  | |  |  | |  |
| Perception of Nutri-Grade labels | 0.91*** | 0.89–0.92 | 0.91*** | 0.89–0.92 | 0.91*** | | 0.89–0.92 | 0.91*** | 0.89–0.92 | 0.91*** | 0.89–0.92 | 0.91*** | | 0.89–0.92 | 0.91*** | | 0.89–0.92 |
| Knowledge of the Nutri-Grade labels | 1.02 | 1.00–1.04 | 1.02 | 1.00–1.04 | 1.02 | | 1.00–1.04 | 1.02 | 1.00–1.04 | 1.02 | 1.00–1.04 | 1.02 | | 1.00–1.04 | 1.02 | | 1.00–1.04 |
| **Built environment** |  |  |  |  |  | |  |  |  |  |  |  | |  |  | |  |
| HDB house price | 0.98 | 0.95–1.01 | 0.97 | 0.94–1.01 | 0.96* | | 0.93–1.00 | 0.96* | 0.93–0.99 | 0.96* | 0.93–1.00 | 0.97 | | 0.94–1.01 | 0.97 | | 0.93–1.00 |
| Commercial land use | 1.05* | 1.01–1.08 | 1.04* | 1.00–1.08 | 1.02 | | 0.99–1.05 | 1.02 | 0.99–1.06 | 1.02 | 0.98–1.06 | 1.04* | | 1.00–1.08 | 1.02 | | 0.98–1.05 |
| Density of Food Courts (per km^2^) |  |  |  |  | 1.02 | | 0.99–1.05 |  |  |  |  |  | |  |  | |  |
| Density of Bars (per km^2^) |  |  |  |  |  | |  | 0.99 | 0.96–1.01 |  |  |  | |  |  | |  |
| Density of Cafe (per km^2^) |  |  |  |  |  | |  |  |  | 1.01 | 0.97–1.04 |  | |  |  | |  |
| Density of vending machines (per km^2^) | 0.96* | 0.93–1.00 | 0.96* | 0.93–1.00 |  | |  |  |  |  |  | 0.96* | | 0.93–1.00 |  | |  |
| Density of juice bars (per km^2^) |  |  |  |  |  | |  |  |  |  |  |  | |  | 1.00 | | 0.97–1.04 |
| **Interaction** |  |  |  |  |  | |  |  |  |  |  |  | |  |  | |  |
| Knowledge of Nutri-Grade labels×HDB house price | 1.05*** | 1.03–1.07 |  |  |  | |  |  |  |  |  |  | |  |  | |  |
| Knowledge of Nutri-Grade labels×Commercial land use |  |  | 1.01 | 0.99–1.03 |  | |  |  |  |  |  |  | |  |  | |  |
| Knowledge of Nutri-Grade labels×Food Courts |  |  |  |  | 1.00 | | 0.98–1.01 |  |  |  |  |  | |  |  | |  |
| Knowledge of Nutri-Grade labels×Bars |  |  |  |  |  | |  | 1.03** | 1.01–1.06 |  |  |  | |  |  | |  |
| Knowledge of Nutri-Grade labels×Cafes |  |  |  |  |  | |  |  |  | 1.02 | 1.00–1.04 |  | |  |  | |  |
| Knowledge of Nutri-Grade labels×Vending machines |  |  |  |  |  | |  |  |  |  |  | 1.01 | | 0.99–1.03 |  | |  |
| Knowledge of Nutri-Grade labels×Juice bars |  |  |  |  |  | |  |  |  |  |  |  | |  | 0.98* | | 0.96–1.00 |
| AIC | 13829.89 |  | 13858.17 |  | 13863.03 | |  | 13854.05 |  | 13861.11 |  | 13858.9 | |  | 13859.18 | |  |
| Log-Likelihood | -6875.95 |  | -6890.08 |  | -6892.52 | |  | -6888.03 |  | -6891.56 |  | -6890.45 | |  | -6890.59 | |  |

IRR: Incident Rate Ratio; CI: Confidence Interval; **p* < .05; ***p* < .01; ****p* < .001.

**Table S4.** Moderation effects of perception of Nutri-Grade labels and built environment factors on the total number of different types of SSBs consumed weekly in Singapore (n = 2,868)

| **Characteristics** | Model 1 | | | Model 2 | | | Model 3 | | | | Model 4 | | | Model 5 | | | | | Model 6 | | | | | Model 7 | | |
| --- | --- | --- | --- | --- | --- | --- | --- | --- | --- | --- | --- | --- | --- | --- | --- | --- | --- | --- | --- | --- | --- | --- | --- | --- | --- | --- |
|  | IRR | 95% CI | IRR | | 95% CI | IRR | | 95% CI | | IRR | | 95% CI | IRR | | 95% CI | | IRR | | | 95% CI | | IRR | | | 95% CI | |
| **Demographic** |  |  |  | |  |  | |  | |  | |  |  | |  | |  | | |  | |  | | |  | |
| **Age** |  |  |  | |  |  | |  | |  | |  |  | |  | |  | | |  | |  | | |  | |
| 21-39 years | Ref |  | Ref | |  | Ref | |  | | Ref | |  | Ref | |  | | Ref | | |  | | Ref | | |  | |
| 40-59 years | 0.91*** | 0.87–0.95 | 0.91*** | | 0.87–0.95 | 0.91*** | | 0.87–0.95 | | 0.91*** | | 0.87–0.95 | 0.91*** | | 0.87–0.95 | | 0.91*** | | | 0.87–0.95 | | 0.91*** | | | 0.87–0.95 | |
| 60 years or above | 0.95 | 0.88–1.02 | 0.94 | | 0.88–1.01 | 0.94 | | 0.88–1.01 | 0.94 | | | 0.88–1.01 | 0.94 | | | 0.88–1.01 | | 0.94 | | | 0.88–1.01 | | 0.94 | | | 0.88–1.01 |
| **Sex** |  |  |  | |  |  | |  |  | | |  |  | | |  | |  | | |  | |  | | |  |
| Male | Ref |  | Ref | |  | Ref | |  | Ref | | |  | Ref | | |  | | Ref | | |  | | Ref | | |  |
| Female | 0.94** | 0.91–0.98 | 0.94** | | 0.91–0.98 | 0.94** | | 0.91–0.98 | 0.94** | | | 0.91–0.98 | 0.94** | | | 0.91–0.98 | | 0.94** | | | 0.91–0.98 | | 0.94** | | | 0.91–0.98 |
| **Ethnicity** |  |  |  | |  |  | |  |  | | |  |  | | |  | |  | | |  | |  | | |  |
| Chinese | Ref |  | Ref | |  | Ref | |  | Ref | | |  | Ref | | |  | | Ref | | |  | | Ref | | |  |
| Malay | 0.89*** | 0.85–0.94 | 0.89*** | | 0.85–0.94 | 0.89*** | | 0.85–0.94 | 0.89*** | | | 0.85–0.94 | 0.89*** | | | 0.85–0.94 | | 0.89*** | | | 0.85–0.94 | | 0.89*** | | | 0.85–0.94 |
| Indian | 0.89*** | 0.84–0.94 | 0.89*** | | 0.84–0.94 | 0.89*** | | 0.84–0.94 | 0.89*** | | | 0.84–0.94 | 0.89*** | | | 0.84–0.94 | | 0.89*** | | | 0.84–0.94 | | 0.89*** | | | 0.84–0.94 |
| Others | 1.11 | 0.95–1.30 | 1.11 | | 0.95–1.30 | 1.11 | | 0.95–1.30 | 1.11 | | | 0.95–1.30 | 1.12 | | | 0.95–1.30 | | 1.11 | | | 0.95–1.30 | | 1.11 | | | 0.95–1.30 |
| **Education level** |  |  |  | |  |  | |  |  | | |  |  | | |  | |  | | |  | |  | | |  |
| Below university | Ref |  | Ref | |  | Ref | |  | Ref | | |  | Ref | | |  | | Ref | | |  | | Ref | | |  |
| University and above | 1.04* | 1.00–1.08 | 1.04* | | 1.00–1.08 | 1.04* | | 1.00–1.08 | 1.04* | | | 1.00–1.08 | 1.04* | | | 1.00–1.08 | | 1.04* | | | 1.00–1.08 | | 1.04* | | | 1.00–1.08 |
| **Marital status** |  |  |  | |  |  | |  |  | | |  |  | | |  | |  | | |  | |  | | |  |
| Married with children | Ref |  | Ref | |  | Ref | |  | Ref | | |  | Ref | | |  | | Ref | | |  | | Ref | | |  |
| Married without children | 1.05 | 0.99–1.11 | 1.05 | | 0.99–1.11 | 1.05 | | 0.99–1.11 | 1.05 | | | 0.99–1.11 | 1.05 | | | 0.99–1.11 | | 1.05 | | | 0.99–1.11 | | 1.05 | | | 0.99–1.11 |
| Single | 0.99 | 0.95–1.04 | 0.99 | | 0.95–1.04 | 0.99 | | 0.94–1.04 | 0.99 | | | 0.94–1.04 | 0.99 | | | 0.94–1.04 | | 0.99 | | | 0.95–1.04 | | 0.99 | | | 0.94–1.04 |
| Widowed/Divorced/Separated | 0.82*** | 0.75–0.90 | 0.82*** | | 0.75–0.90 | 0.82*** | | 0.75–0.90 | 0.82*** | | | 0.75–0.90 | 0.82*** | | | 0.75–0.90 | | 0.82*** | | | 0.75–0.90 | | 0.82*** | | | 0.75–0.90 |
| **Employment status** |  |  |  | |  |  | |  |  | | |  |  | | |  | |  | | |  | |  | | |  |
| Employed full-time | Ref |  | Ref | |  | Ref | |  | Ref | | |  | Ref | | |  | | Ref | | |  | | Ref | | |  |
| Employed part-time | 1.00 | 0.93–1.08 | 1.00 | | 0.93–1.08 | 1.00 | | 0.93–1.08 | 1.00 | | | 0.93–1.08 | 1.00 | | | 0.93–1.08 | | 1.00 | | | 0.93–1.08 | | 1.00 | | | 0.93–1.08 |
| Others | 0.98 | 0.86–1.11 | 0.98 | | 0.86–1.11 | 0.98 | | 0.86–1.12 | 0.98 | | | 0.86–1.11 | 0.98 | | | 0.86–1.11 | | 0.98 | | | 0.86–1.11 | | 0.98 | | | 0.86–1.11 |
| Retired/Homemaker | 1.02 | 0.96–1.09 | 1.02 | | 0.96–1.09 | 1.02 | | 0.96–1.09 | 1.02 | | | 0.96–1.09 | 1.02 | | | 0.96–1.09 | | 1.02 | | | 0.96–1.09 | | 1.02 | | | 0.96–1.09 |
| Self-employed | 0.86** | 0.78–0.94 | 0.86** | | 0.79–0.94 | 0.86*** | | 0.78–0.94 | 0.86** | | | 0.79–0.94 | 0.86** | | | 0.78–0.94 | | 0.86** | | | 0.79–0.94 | | 0.86** | | | 0.78–0.94 |
| Unemployed | 1.06 | 0.95–1.19 | 1.07 | | 0.95–1.19 | 1.07 | | 0.95–1.19 | 1.07 | | | 0.95–1.19 | 1.07 | | | 0.95–1.19 | | 1.06 | | | 0.95–1.19 | | 1.07 | | | 0.95–1.19 |
| **Health status** |  |  |  | |  |  | |  |  | | |  |  | | |  | |  | | |  | |  | | |  |
| **BMI** |  |  |  | |  |  | |  |  | | |  |  | | |  | |  | | |  | |  | | |  |
| Normal 18.5-22.9 | Ref |  | Ref | |  | Ref | |  | Ref | | |  | Ref | | |  | | Ref | | |  | | Ref | | |  |
| Underweight <18.5 | 1.02 | 0.95–1.09 | 1.02 | | 0.95–1.09 | 1.02 | | 0.95–1.10 | 1.02 | | | 0.95–1.09 | 1.02 | | | 0.95–1.09 | | 1.02 | | | 0.95–1.09 | | 1.02 | | | 0.95–1.09 |
| Overweight 23-27.4 | 1.00 | 0.96–1.04 | 1.00 | | 0.96–1.04 | 1.00 | | 0.96–1.04 | 1.00 | | | 0.96–1.04 | 1.00 | | | 0.96–1.04 | | 1.00 | | | 0.96–1.04 | | 1.00 | | | 0.96–1.04 |
| Obese ≥27.5 | 1.08* | 1.02–1.14 | 1.08* | | 1.02–1.14 | 1.08* | | 1.02–1.14 | 1.08* | | | 1.02–1.14 | 1.08* | | | 1.02–1.14 | | 1.08* | | | 1.02–1.14 | | 1.08* | | | 1.02–1.14 |
| **Medical conditions** |  |  |  | |  |  | |  |  | | |  |  | | |  | |  | | |  | |  | | |  |
| Yes | Ref |  | Ref | |  | Ref | |  | Ref | | |  | Ref | | |  | | Ref | | |  | | Ref | | |  |
| No | 1.05* | 1.01–1.09 | 1.05* | | 1.00–1.09 | 1.04* | | 1.00–1.09 | 1.04* | | | 1.00–1.09 | 1.04* | | | 1.00–1.09 | | 1.05* | | | 1.00–1.09 | | 1.04* | | | 1.00–1.09 |
| **Individuals’ knowledge and belief** |  |  |  | |  |  | |  |  | | |  |  | | |  | |  | | |  | |  | | |  |
| Knowledge of SSBs | 0.96*** | 0.94–0.98 | 0.96*** | | 0.94–0.98 | 0.96*** | | 0.94–0.98 | 0.96*** | | | 0.94–0.98 | 0.96*** | | | 0.94–0.98 | | 0.96*** | | | 0.94–0.98 | | 0.96*** | | | 0.94–0.98 |
| Perceived control of SSB intake | 0.97* | 0.95–1.00 | 0.97* | | 0.95–1.00 | 0.97 | | 0.95–1.00* | 0.97* | | | 0.95–1.00 | 0.97* | | | 0.95–1.00 | | 0.97* | | | 0.95–1.00 | | 0.97* | | | 0.95–1.00 |
| Self-efficacy | 0.97** | 0.94–0.99 | 0.97** | | 0.94–0.99 | 0.97** | | 0.94–0.99 | 0.97** | | | 0.94–0.99 | 0.97** | | | 0.94–0.99 | | 0.97** | | | 0.94–0.99 | | 0.97** | | | 0.94–0.99 |
| **Physical environment** |  |  |  | |  |  | |  |  | | |  |  | | |  | |  | | |  | |  | | |  |
| Household availability of SSBs |  |  |  | |  |  | |  |  | | |  |  | | |  | |  | | |  | |  | | |  |
| Very rarely | Ref |  | Ref | |  | Ref | |  | Ref | | |  | Ref | | |  | | Ref | | |  | | Ref | | |  |
| Monthly | 1.51*** | 1.40–1.63 | 1.51*** | | 1.39–1.63 | 1.51*** | | 1.39–1.63 | 1.51*** | | | 1.40–1.63 | 1.51*** | | | 1.39–1.63 | | 1.51*** | | | 1.39–1.63 | | 1.51*** | | | 1.39–1.63 |
| Weekly | 1.59*** | 1.47–1.72 | 1.59*** | | 1.47–1.72 | 1.58*** | | 1.46–1.71 | 1.59*** | | | 1.47–1.72 | 1.58*** | | | 1.46–1.71 | | 1.59*** | | | 1.47–1.72 | | 1.58*** | | | 1.46–1.71 |
| Every other day/daily | 1.62*** | 1.50–1.75 | 1.62*** | | 1.50–1.75 | 1.61*** | | 1.49–1.74 | 1.62*** | | | 1.50–1.75 | 1.61*** | | | 1.49–1.74 | | 1.62*** | | | 1.50–1.75 | | 1.61*** | | | 1.49–1.74 |
| Purchase of SSBs via online delivery |  |  |  | |  |  | |  |  | | |  |  | | |  | |  | | |  | |  | | |  |
| Very rarely | Ref |  | Ref | |  | Ref | |  | Ref | | |  | Ref | | |  | | Ref | | |  | | Ref | | |  |
| Monthly | 1.43*** | 1.35–1.52 | 1.43*** | | 1.35–1.52 | 1.44*** | | 1.36–1.53 | 1.43*** | | | 1.35–1.52 | 1.44*** | | | 1.36–1.52 | | 1.43*** | | | 1.35–1.52 | | 1.44*** | | | 1.36–1.52 |
| Weekly | 1.49*** | 1.41–1.58 | 1.49*** | | 1.40–1.58 | 1.50*** | | 1.41–1.59 | 1.49*** | | | 1.41–1.58 | 1.50*** | | | 1.41–1.59 | | 1.49*** | | | 1.40–1.58 | | 1.50*** | | | 1.41–1.59 |
| Every other day/daily | 1.37*** | 1.30–1.45 | 1.37*** | | 1.30–1.45 | 1.38*** | | 1.30–1.46 | 1.37*** | | | 1.30–1.45 | 1.38*** | | | 1.30–1.46 | | 1.37*** | | | 1.29–1.45 | | 1.38*** | | | 1.30–1.46 |
| **Social environment** |  |  |  | |  |  | |  |  | | |  |  | | |  | |  | | |  | |  | | |  |
| Perceived social norm of acceptance of SSB intake | 1.08*** | 1.06–1.11 | 1.08*** | | 1.06–1.11 | 1.08*** | | 1.06–1.11 | 1.08*** | | | 1.06–1.11 | 1.08*** | | | 1.06–1.11 | | 1.08*** | | | 1.06–1.11 | | 1.08*** | | | 1.06–1.11 |
| Exposure to SSB advertising | 1.05*** | 1.03–1.08 | 1.05*** | | 1.03–1.08 | 1.05*** | | 1.03–1.08 | 1.05*** | | | 1.03–1.08 | 1.05*** | | | 1.03–1.08 | | 1.05*** | | | 1.03–1.08 | | 1.05*** | | | 1.03–1.08 |
| **Nutri-Grade FOPL influences** |  |  |  | |  |  | |  |  | | |  |  | | |  | |  | | |  | |  | | |  |
| Perception of Nutri-Grade labels | 0.91*** | 0.89–0.92 | 0.91*** | | 0.89–0.92 | 0.91*** | | 0.89–0.92 | 0.91*** | | | 0.89–0.92 | 0.91*** | | | 0.89–0.92 | | 0.91*** | | | 0.89–0.92 | | 0.91*** | | | 0.89–0.92 |
| Knowledge of the Nutri-Grade labels | 1.02 | 1.00–1.04 | 1.02 | | 1.00–1.04 | 1.02 | | 1.00–1.04 | 1.02 | | | 1.00–1.04 | 1.02 | | | 1.00–1.04 | | 1.02 | | | 1.00–1.04 | | 1.02 | | | 1.00–1.04 |
| **Built environment** |  |  |  | |  |  | |  |  | | |  |  | | |  | |  | | |  | |  | | |  |
| HDB house price | 0.97 | 0.94–1.00 | 0.97 | | 0.94–1.00 | 0.96* | | 0.93–1.00 | 0.96* | | | 0.92–0.99 | 0.96* | | | 0.93–1.00 | | 0.97 | | | 0.94–1.00 | | 0.96 | | | 0.93–1.00 |
| Commercial land use | 1.04* | 1.00–1.08 | 1.04* | | 1.00–1.08 | 1.02 | | 0.99–1.05 | 1.02 | | | 0.99–1.06 | 1.02 | | | 0.98–1.05 | | 1.04* | | | 1.00–1.08 | | 1.02 | | | 0.98–1.05 |
| Density of Food Courts (per km^2^) |  |  |  | |  | 1.02 | | 0.99–1.05 |  | | |  |  | | |  | |  | | |  | |  | | |  |
| Density of Bars (per km^2^) |  |  |  | |  |  | |  | 0.98 | | | 0.95–1.00 |  | | |  | |  | | |  | |  | | |  |
| Density of Cafe (per km^2^) |  |  |  | |  |  | |  |  | | |  | 1.00 | | | 0.97–1.04 | |  | | |  | |  | | |  |
| Density of vending machines (per km^2^) | 0.96* | 0.93–1.00 | 0.96* | | 0.93–1.00 |  | |  |  | | |  |  | | |  | | 0.96* | | | 0.93–1.00 | |  | | |  |
| Density of juice bars (per km^2^) |  |  |  | |  |  | |  |  | | |  |  | | |  | |  | | |  | | 1.00 | | | 0.97–1.04 |
| **Interaction** |  |  |  | |  |  | |  |  | | |  |  | | |  | |  | | |  | |  | | |  |
| Perception of Nutri-Grade labels×HDB house price | 0.99 | 0.97–1.01 |  | |  |  | |  |  | | |  |  | | |  | |  | | |  | |  | | |  |
| Perception of Nutri-Grade labels×Commercial land use |  |  | 1.00 | | 0.98–1.02 |  | |  |  | | |  |  | | |  | |  | | |  | |  | | |  |
| Perception of Nutri-Grade labels×Food Courts |  |  |  | |  | 1.01 | | 0.99–1.02 |  | | |  |  | | |  | |  | | |  | |  | | |  |
| Perception of Nutri-Grade labels×Bars |  |  |  | |  |  | |  | 1.00 | | | 0.98–1.02 |  | | |  | |  | | |  | |  | | |  |
| Perception of Nutri-Grade labels×Cafes |  |  |  | |  |  | |  |  | | |  | 1.00 | | | 0.98–1.02 | |  | | |  | |  | | |  |
| Perception of Nutri-Grade labels×Vending machines |  |  |  | |  |  | |  |  | | |  |  | | |  | | 1.00 | | | 0.98–1.02 | |  | | |  |
| Perception of Nutri-Grade labels×juice bars |  |  |  | |  |  | |  |  | | |  |  | | |  | |  | | |  | | 1.00 | | | 0.98–1.02 |
| AIC | 13859.65 |  | 13860.17 | |  | 13862.78 | |  | 13861.37 | | |  | 13864.57 | | |  | | 13860.0 | | |  | | 13864.53 | | |  |
| Log-Likelihood | -6890.83 |  | -6891.09 | |  | -6892.39 | |  | -6891.68 | | |  | -6893.28 | | |  | | -6891.0 | | |  | | -6893.26 | | |  |

IRR: Incident Rate Ratio; CI: Confidence Interval; **p* < .05; ***p* < .01; ****p* < .001.

**Table S5.** Moderation effects of age groups with knowledge and perception of Nutri-Grade labels in Singapore (n = 2,868)

| **Characteristics** | SSB total intake/times | | | | Total SSB types consumed/counts | | | |
| --- | --- | --- | --- | --- | --- | --- | --- | --- |
|  | Model 1 | | Model 2 | | Model 1 | | Model 2 | |
|  | AOR | 95% CI | AOR | 95% CI | IRR | 95% CI | IRR | 95% CI |
| **Demographic** |  |  |  |  |  |  |  |  |
| **Age** |  |  |  |  |  |  |  |  |
| 21-39 years | Ref |  | Ref |  | Ref |  | Ref |  |
| 40-59 years | 0.80* | 0.67–0.96 | 0.80* | 0.67–0.96 | 0.91*** | 0.87–0.95 | 0.91*** | 0.87–0.95 |
| 60 years or above | 1.45* | 1.07–1.97 | 1.45* | 1.07–1.96 | 0.94 | 0.88–1.01 | 0.94 | 0.88–1.01 |
| **Sex** |  |  |  |  |  |  |  |  |
| Male | Ref |  | Ref |  | Ref |  | Ref |  |
| Female | 0.79** | 0.68–0.91 | 0.78*** | 0.67–0.90 | 0.95** | 0.91–0.98 | 0.94** | 0.91–0.98 |
| **Ethnicity** |  |  |  |  |  |  |  |  |
| Chinese | Ref |  | Ref |  | Ref |  | Ref |  |
| Malay | 0.75** | 0.61–0.92 | 0.75** | 0.61–0.92 | 0.89*** | 0.85–0.94 | 0.89*** | 0.85–0.94 |
| Indian | 0.97 | 0.78–1.22 | 0.96 | 0.77–1.20 | 0.90*** | 0.85–0.95 | 0.89*** | 0.84–0.94 |
| Others | 2.63** | 1.31–5.28 | 2.64** | 1.32–5.31 | 1.11 | 0.95–1.29 | 1.11 | 0.95–1.30 |
| **Education level** |  |  |  |  |  |  |  |  |
| Below university | Ref |  | Ref |  | Ref |  | Ref |  |
| University and above | 0.97 | 0.83–1.13 | 0.97 | 0.83–1.13 | 1.04* | 1.01–1.08 | 1.04* | 1.00–1.08 |
| **Marital status** |  |  |  |  |  |  |  |  |
| Married with children | Ref |  | Ref |  | Ref |  | Ref |  |
| Married without children | 0.92 | 0.72–1.17 | 0.93 | 0.73–1.18 | 1.05 | 0.99–1.11 | 1.05 | 0.99–1.11 |
| Single | 0.82 | 0.68–1.00 | 0.84 | 0.69–1.02 | 0.99 | 0.94–1.04 | 0.99 | 0.94–1.04 |
| Widowed/Divorced/Separated | 0.44*** | 0.32–0.61 | 0.44*** | 0.32–0.61 | 0.82*** | 0.75–0.90 | 0.82*** | 0.75–0.90 |
| **Employment status** |  |  |  |  |  |  |  |  |
| Employed full-time | Ref |  | Ref |  | Ref |  | Ref |  |
| Employed part-time | 1.10 | 0.81–1.47 | 1.10 | 0.82–1.48 | 1.00 | 0.93–1.08 | 1.00 | 0.93–1.08 |
| Others | 0.72 | 0.42–1.22 | 0.73 | 0.43–1.24 | 0.98 | 0.86–1.11 | 0.98 | 0.86–1.11 |
| Retired/Homemaker | 1.07 | 0.82–1.38 | 1.08 | 0.83–1.39 | 1.02 | 0.96–1.09 | 1.02 | 0.96–1.09 |
| Self-employed | 0.63** | 0.45–0.88 | 0.63** | 0.45–0.88 | 0.86** | 0.79–0.94 | 0.86** | 0.79–0.94 |
| Unemployed | 1.41 | 0.89–2.23 | 1.38 | 0.87–2.19 | 1.07 | 0.95–1.19 | 1.07 | 0.95–1.19 |
| **Health status** |  |  |  |  |  |  |  |  |
| **BMI** |  |  |  |  |  |  |  |  |
| Normal 18.5-22.9 | Ref |  | Ref |  | Ref |  | Ref |  |
| Underweight <18.5 | 0.96 | 0.71–1.28 | 0.95 | 0.71–1.28 | 1.02 | 0.95–1.10 | 1.02 | 0.95–1.09 |
| Overweight 23-27.4 | 0.82* | 0.70–0.97 | 0.81* | 0.69–0.96 | 1.00 | 0.96–1.04 | 1.00 | 0.96–1.04 |
| Obese ≥27.5 | 1.15 | 0.90–1.46 | 1.14 | 0.90–1.46 | 1.08* | 1.02–1.14 | 1.08* | 1.02–1.14 |
| **Medical conditions** |  |  |  |  |  |  |  |  |
| Yes | Ref |  | Ref |  | Ref |  | Ref |  |
| No | 1.29** | 1.10–1.52 | 1.29** | 1.10–1.52 | 1.05* | 1.01–1.09 | 1.04* | 1.00–1.09 |
| **Individuals’ knowledge and belief** |  |  |  |  |  |  |  |  |
| Knowledge of SSBs | 0.85*** | 0.78–0.92 | 0.84*** | 0.77–0.91 | 0.96*** | 0.94–0.98 | 0.96*** | 0.94–0.98 |
| Perceived control of SSB intake | 0.81*** | 0.74–0.90 | 0.81*** | 0.74–0.90 | 0.97* | 0.95–1.00 | 0.97* | 0.95–1.00 |
| Self-efficacy | 0.97 | 0.88–1.07 | 0.97 | 0.88–1.07 | 0.97** | 0.94–0.99 | 0.97** | 0.94–0.99 |
| **Physical environment** |  |  |  |  |  |  |  |  |
| Household availability of SSBs |  |  |  |  |  |  |  |  |
| Very rarely | Ref |  | Ref |  | Ref |  | Ref |  |
| Monthly | 3.06*** | 2.32–4.04 | 3.08*** | 2.33–4.07 | 1.51*** | 1.39–1.63 | 1.51*** | 1.39–1.63 |
| Weekly | 3.62*** | 2.70–4.85 | 3.67*** | 2.74–4.92 | 1.58*** | 1.46–1.71 | 1.59*** | 1.47–1.72 |
| Every other day/daily | 4.31*** | 3.23–5.74 | 4.40*** | 3.31–5.86 | 1.61*** | 1.49–1.74 | 1.62*** | 1.49–1.75 |
| Purchase of SSBs via online delivery |  |  |  |  |  |  |  |  |
| Very rarely | Ref |  | Ref |  | Ref |  | Ref |  |
| Monthly | 2.65*** | 2.10–3.34 | 2.58*** | 2.05–3.25 | 1.44*** | 1.36–1.52 | 1.43*** | 1.35–1.52 |
| Weekly | 2.98*** | 2.33–3.80 | 2.92*** | 2.29–3.72 | 1.50*** | 1.41–1.59 | 1.49*** | 1.41–1.58 |
| Every other day/daily | 2.44*** | 1.95–3.06 | 2.42*** | 1.93–3.04 | 1.37*** | 1.30–1.46 | 1.37*** | 1.29–1.45 |
| **Social environment** |  |  |  |  |  |  |  |  |
| Perceived social norm of acceptance of SSB intake | 1.53*** | 1.40–1.68 | 1.52*** | 1.39–1.67 | 1.09*** | 1.06–1.11 | 1.09*** | 1.06–1.11 |
| Exposure to SSB advertising | 1.23*** | 1.12–1.35 | 1.24*** | 1.13–1.36 | 1.05*** | 1.03–1.08 | 1.05*** | 1.03–1.08 |
| **Nutri-Grade FOPL influences** |  |  |  |  |  |  |  |  |
| Perception of Nutri-Grade labels | 0.72*** | 0.67–0.78 | 0.69*** | 0.61–0.78 | 0.91*** | 0.89–0.92 | 0.91*** | 0.89–0.94 |
| Knowledge of the Nutri-Grade labels | 1.17* | 1.03–1.34 | 1.04 | 0.96–1.13 | 1.04* | 1.01–1.07 | 1.02 | 1.00–1.04 |
| **Built environment** |  |  |  |  |  |  |  |  |
| HDB house price | 0.84* | 0.73–0.96 | 0.84* | 0.73–0.97 | 0.97 | 0.94–1.00 | 0.97 | 0.94–1.00 |
| Commercial land use | 1.12 | 0.98–1.28 | 1.13 | 0.99–1.29 | 1.04* | 1.00–1.08 | 1.04* | 1.00–1.08 |
| Density of Food Courts (per km^2^) | 1.08 | 0.95–1.23 | 1.09 | 0.95–1.24 |  |  |  |  |
| Density of vending machines (per km^2^) |  |  |  |  | 0.96* | 0.93–0.99 | 0.96* | 0.93–1.00 |
| **Interaction** |  |  |  |  |  |  |  |  |
| Knowledge of Nutri-Grade labels×age 40-59 (reference group: age 21-30) | 0.81* | 0.69–0.95 |  |  | 0.96 | 0.93–1.00 |  |  |
| Knowledge of Nutri-Grade labels×age 60 or above (reference group: age 21-30) | 0.92 | 0.72–1.18 |  |  | 0.97 | 0.91–1.03 |  |  |
| Perception of Nutri-Grade labels×age 40-59 (reference group: age 21-30) |  |  | 1.08 | 0.93–1.26 |  |  | 0.99 | 0.95–1.02 |
| Perception of Nutri-Grade labels×age 60 or above (reference group: age 21-30) |  |  | 1.00 | 0.78–1.28 |  |  | 0.99 | 0.93–1.05 |
| AIC | 6942.06 |  | 6947.59 |  | 13858.47 |  | 13861.72 |  |
| Log-Likelihood | -3429.03 |  | -3431.79 |  | -6889.24 |  | -6890.86 |  |

AOR: Adjusted Odds Ratio; CI: Confidence Interval; IRR: Incident Rate Ratio; **p* < .05; ***p* < .01; ****p* < .001.
